# Characterising contact in disease outbreaks via a network model of spatial-temporal proximity

**DOI:** 10.1101/2021.04.07.21254497

**Authors:** Ashleigh Myall, Robert L. Peach, Yu Wan, Siddharth Mookerjee, Elita Jauneikaite, Frankie Bolt, James Price, Frances Davies, Andrea Y. Weiße, Alison Holmes, Mauricio Barahona

**Affiliations:** Department of Mathematics, Imperial College London, London, UK; NIHR Health Protection Research Unit in Healthcare Associated Infections and Antimicrobial Resistance, Department of Infectious Disease, Imperial College London, London, UK; Department of Neurology, University Hospital Würzburg, Würzburg, Germany; Department of Brain Sciences, Imperial College London, London, UK; Imperial College Healthcare NHS Trust, London, UK; Department of Infectious Disease Epidemiology, School of Public Health, Imperial College London, London, UK; School of Informatics, University of Edinburgh, Scotland, UK; School of Biological Sciences, University of Edinburgh, Scotland, UK

## Abstract

Contact tracing is a key tool in epidemiology to identify and control outbreaks of infectious diseases. Existing contact tracing methodologies produce contact maps of individuals based on a binary definition of contact which can be hampered by missing data and indirect contacts. Here, we present a Spatial-temporal Epidemiological Proximity (StEP) model to recover contact maps in disease outbreaks based on movement data. The StEP model accounts for imperfect data by considering probabilistic contacts between individuals based on spatial-temporal proximity of their movement trajectories, creating a robust movement network despite possible missing data and unseen transmission routes. Using real-world data we showcase the potential of StEP for contact tracing with outbreaks of multidrug-resistant bacteria and COVID-19 in a large hospital group in London, UK. In addition to the core structure of contacts that can be recovered using traditional methods of contact tracing, the StEP model reveals missing contacts that connect seemingly separate outbreaks. Comparison with genomic data further confirmed that these recovered contacts indeed improve characterisation of disease transmission and so highlights how the StEP framework can inform effective strategies of infection control and prevention.

## Main

The spread of infectious diseases, direct or indirect, is predominantly mediated by person-to-person contacts^1^. Establishment of contacts, otherwise known as contact tracing, aims to uncover such transmission events and provide a basis to direct disease control policies^2^. Contact tracing plays a pivotal role in the public health response to many diseases. For example, it guided targeted vaccination that led to the eradication of smallpox in many areas^3^. For HIV, contact tracing provided the first evidence that its spread was primarily transmitted through sexual contact, which subsequently led to targeted safe-sex campaigns^4–7^. It also proved an effective public health tool when it helped limit the 2003 outbreak of SARS in China^8^. Recently, contact tracing has been a key component of the response to COVID-19^9^, where countries and areas that aggressively traced contacts of confirmed cases, such as Singapore, South Korea, New Zealand, mainland China, and Taiwan, have all maintained low case rates which can be attributed largely to their prompt tracking and isolation of spreaders^10–12^.

Contact tracing establishes routes of disease transmission based on a network of contacts between affected individuals^13^. Its effectiveness highly depends on both the quality of data and how a ‘contact’ is defined, for example, a contact may be established when individuals share a location over a certain period of time. Such discrete definition, however, can miss certain transmission events^14–18^, and unidentified individuals as well as contaminated environments can moreover contribute to *indirect* contacts that lead to transmission^19^ but are overlooked by contact tracing. Indirect or unseen pathways of transmission through intermediate contacts result in missing contacts between known infected individuals (Figure 1), which inevitably hamper investigations and can misguide conclusions^20,21^.

**Figure 1.**
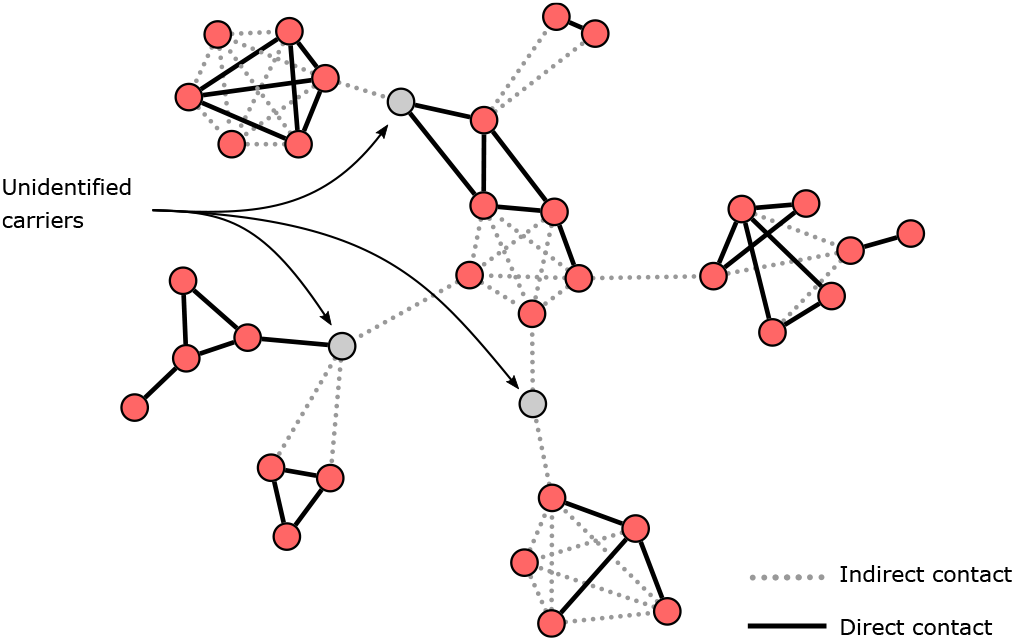
Contact network between individuals in a disease outbreak investigation. Establishing networks of contact is frequently used in disease outbreak investigations. However, it can be hampered by missing data, for example, when contacts between carriers are unobserved or non-observable, or when contact is mediated by unknown carriers and environmental surfaces. Ignoring these routes when investigating outbreaks and transmission routes can lead to a mis-characterisation of disease outbreaks.

Mobility patterns of individuals within a system are a primary vector for disease spread^22–25^. From the Black Death sweeping across medieval Europe via inter-town population movements^26^ to the 1918 Flu Pandemic which followed troop movements in its early stage^27^, mobility data has proven effective in explaining spatial disease dynamics. More recently, flight data has been used for accurate prediction of global disease spread^28–30^, and inter-hospital spread of disease has been shown to closely follow patient referrals^31^. Taking movement flows into account, moreover, allows recovery of missing transmission events, as it quantifies how connected two individuals have been over time and space, and thus, how likely transmission occurred.

Here, we present the Spatial-temporal Epidemiological Proximity (StEP) model, which assumes that the likelihood of transmission between individuals given their spatial-temporal (space and time) proximity. StEP assumes that the likelihood of transmission scales with the spatial-temporal proximity of the individuals’ time-stamped trajectories, defined via a parametric kernel function. Two types of data are taken as input: (1) the movement trajectories of confirmed cases; (2) information on the total movement flows of all individuals in the system, irrespective of their infection status. To quantify the likelihood of transmission among positive cases, StEP relies on two parameters that can be trained to reflect epidemiological characteristics of a given pathogen. The output is a contact network where nodes represent identified cases and edges plausible transmission routes between them. We have implemented our methods in an R package StEP (github.com/ashm97/StEP).

We demonstrate the effectiveness of StEP in three case studies covering in total five hospital-acquired disease outbreaks. Four of the data sets comprise patients colonised with carbapenemase-producing *Enterobacteriaceae* (CPE), a multidrug-resistant pathogen with a high propensity to spread in healthcare settings^32,33^. In our first case study, we trained and assessed StEP on an outbreak of a specific type of CPE, CPE_IMP_^34^, identified in 116 patients over three years. We used semi-supervised learning to train parameters that align the resulting contact network with the distribution of transmission markers (approximating overall transmission structure) available for a subset of confirmed cases. To assess transmission alignment, we then compared the contact network to a whole-genome sequencing (WGS) analysis of a plasmid identified in most CPE_IMP_ isolates carrying several acquired resistance genes ^35^. In our second case study, we transfer the optimised CPE_IMP_ model to analyse further three CPE outbreaks, demonstrating that once trained, StEP can be deployed for a similar class of pathogen without the need for additional labelled data. The efficacy of this approach was validated using less accurate but routinely collected transmission markers. Finally, in our third case study, we analyse data from a hospital outbreak of COVID-19. At the time, COVID-19 was a new pathogen. Since we lacked labelled data to track transmission, we explored StEP’s efficacy for unsupervised learning, by aligning the contact networks to topological signals identified in the unsupervised investigation across previous case studies.

## Spatial-temporal epidemiological proximity (StEP) model

The base assumption of the StEP model, summarised in Figure 2A, is that disease transmission is likely to occur between the most connected physical locations^26^. Connectivity between locations is quantified by the total movement of all individuals in a system and represented by an effective distance network among locations^36^. A pairwise proximity between infected individuals can be computed based on their movement histories across the effective distance network. The resulting contact map quantifies the likelihood of disease transmission between known cases via direct and indirect contacts.

**Figure 2.**
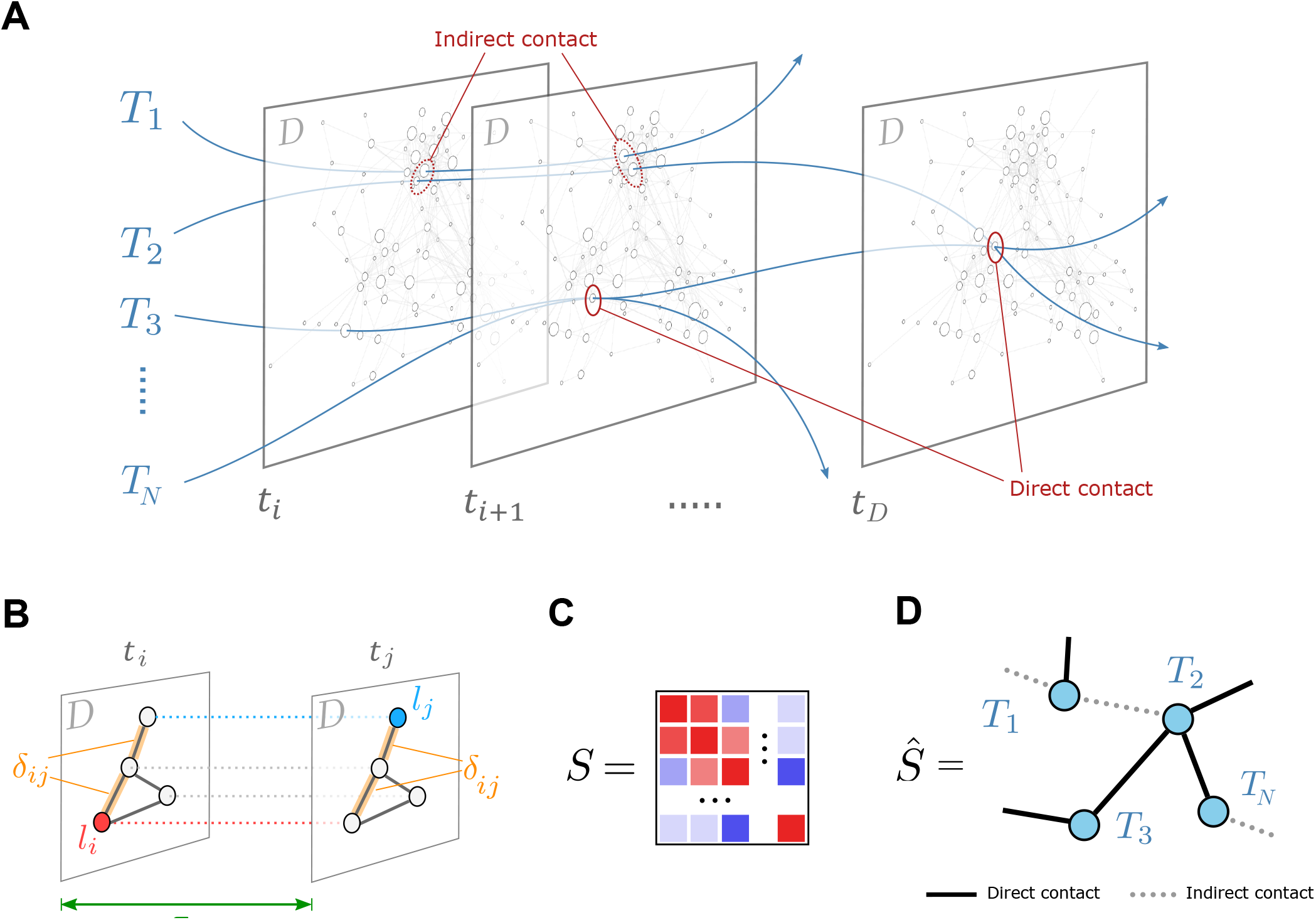
Overview of the StEP framework. **(A)** Movement histories of infected individuals are represented by a set of network trajectories {*T*_1_, *T*_2_, *T*_3_, …, *T*_*N*_} transiently passing through the physical locations of the background movement network *D*. (B) The model accounts for direct and indirect contacts by measuring the distance between time-stamped locations in terms of their spatial proximity *δ*_*ij*_ (with respect to the effective distance network D) and temporal proximity *τ*_*ij*_. (C) The *N* ×*N* similarity matrix *Ŝ* quantifies the total spatial-temporal proximity between any two patient trajectories. (D) The filtered similarity matrix *Ŝ* contains only the strongest spatial-temporal contacts to reveal most likely routes of transmission.

### Contacts mediated by total movement

We consider an effective distance network *D* = (*V, E*) that is derived from the total movement of individuals in a system (Figure S1). The nodes *V* = {*v*_*i*_} correspond to physical locations and the edges *E*_*i j*_ = {*w*_*i j*_} are weighted by the effective distance of total movement between locations *v*_*i*_ → *v* _*j*_^36^ (see Methods: Effective distance). Effective distance is a probabilistic measure of likely disease pathways and has been shown to capture disease propagation across real-world mobility networks^36^. By mapping locations visited by infected individuals onto the effective distance matrix of the hospital network we are able to quantify the spatial-temporal proximity and recover contact structures underpinning disease propagation.

### Spatial-temporal proximity between trajectories

We consider the movement histories of *N* infected individuals under investigation represented by a set of trajectories *𝒯* = {*T*_1_, *T*_2_, *T*_3_, …, *T*_*N*_}. Each trajectory *T*_*n*_ is an ordered set of location-timings

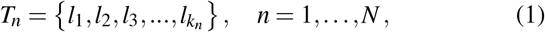

where *k*_*n*_ is the number of location-timings for individual *n*, and each location-timing *l*_*i*_ = (*v*_*i*_, *t*_*i*_) is a tuple of the ward location *v*_*i*_ ∈*V* visited by the individual at time *t*_*i*_. Rather than considering only direct contacts, where trajectories overlap *T*_*n*_ ∩*T*_*m*_*≠* ø, we consider the likelihood of transmission given the proximity of trajectories in space and time relative to the background disease propagation.

We define the spatial-temporal proximity between location-timings *l*_*i*_ and *l* _*j*_ via a kernel

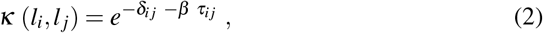

where *τ*_*i j*_ = |*t*_*i*_ −*t* _*j*_ |is the absolute time difference and *δ*_*i j*_ the effective distance between wards *v*_*i*_ and *v* _*j*_ (Figure 2B) defined by the shortest-path distance (i.e. the most likely path of disease transmission) between wards *v*_*i*_ and *v* _*j*_ on the distance network *D* (see Methods: Effective distance). The parameter *β* represents the propagation speed of the disease and can be adpated to the pathogen under investigation.

Based on the proximity between location-timings (2), we then compute the total proximity between two trajectories *T*_*m*_ and *T*_*n*_ by summing over the pairwise proximities of locationtimings

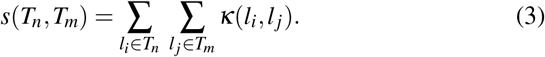

The total proximity *s* scales quadratically with the lengths of trajectories, reflecting the increased likelihood of a direct or indirect transmission when two patients coincide for longer periods^37^.Finally, the trajectory-to-trajectory (*N* × *N*) matrix *S* with elements *S*_*nm*_ = *s*(*T*_*n*_, *T*_*m*_) summarises the Spatialtemporal proximity among individual movement histories (Figure 2C).

### Filtering contacts to reveal likely transmission

To reveal the most likely transmission routes, we filter out weak connections from the similarity matrix *S*. We use an extension to *k*-nearest neighbours called continuous *k*-nearest neighbours (CkNN)^39^ for edge filtering which performs well topological features span several different scales^40^, as is the case for disease outbreaks spanning across different scales in time and space (see Methods: CkNN). The filtered similarity matrix *Ŝ* then includes up to *k* of the strongest contacts for each individual. We refer to the input parameter *k* to CkNN as the edge density. Edge density reflects pathogen infectiousness, e.g. pathogens that transmit via minimal spatial-temporal contact require high values of *k*. The resulting matrix *S* can be interpreted as the adjacency matrix of a contact network, where each node corresponds to an infected individual (Figure 2D) and edges to contacts and thus possible transmission events^13^.

### Model training

The two parameters of StEP, the propagation speed *β* and the edge density *k*, reflect pathogen-specific transmission dynamics. In our case studies, we present three different approaches to parameter learning. Firstly, we use graph (semi-)supervised learning to predict node labels on genomic markers and identify the contact network that maximises the classification accuracy (see Methods: Semi-supervised learning of parameters). The semi-supervised approach enables learning from partially labelled data via label diffusion to model transmission across the contact network. Secondly, we transfer the previously trained model to new pathogens that are epidemiologically similar and studied in the same environment, thus avoiding the use of additional labelled data. Thirdly, we use StEP entirely unsupervised to investigate an outbreak of an unrelated pathogen without ground-truth labels available, exploiting structural statistics of the contact networks to tune parameters (see Methods: Unsupervised learning of parameters) and validating outcomes on data where optimal parameters are known.

## Results

### Case study 1: Outbreak of CPE_IMP_

CPE isolates carrying the imipenem-resistance gene *bla*_IMP_, denoted as CPE_IMP_, were rare and sporadic in the UK as of 2015^41^. Following a seeding case in 2016 in our hospital group of study, however, within trust reports of CPE_IMP_ incidence increased, and over the subsequent three years were identified in 116 hospital patients^35^. To understand CPE_IMP_s spread, 85 isolates from confirmed CPE_IMP_-carrying patients were sampled for WGS and analysed. The WGS analysis results were used to construct 181 biomarkers based on the presence of certain acquired resistance genes alleles (see Methods: Biomarker construction), enabling transmission tracing. We used these biomarkers together with patient trajectories as input for StEP to learn epidemiological parameters specific to CPE_IMP_ and construct the patient contact network.

### Learning pathogen-specific parameters

We used Graph Diffusion Reclassification (GDR)^42^ to diffuse labels (biomarkers in this study) over the complete network and quantified accuracy in predicting biomarkers by the *F*_1_-score (Figure 3A). Prediction accuracy is maximal for propagation speed *β* = 0.6 and edge density *k* = 3, suggesting that the resulting contact network best aligns with transmission of CPE_IMP_. In this opti-mal region, we observe substantial heterogeneity across the performance of individual biomarkers, many of which seem unrelated to outbreaks (Table s1). Among the 181 biomarkers, 51 reached mean *F*_1_-scores above 0.6, and in particular, nine biomarkers performed particularly well, reaching a mean *F*_1_-score above 0.9. The latter nine biomarkers were also strongly associated with plasmid clusters independently identified through genomic analysis^35^ (Figure S2). Hence, it is likely the diffusion of biomarkers over StEP’s recovered contact network is well characterising parts of the CPE_IMP_ outbreak.

**Figure 3.**
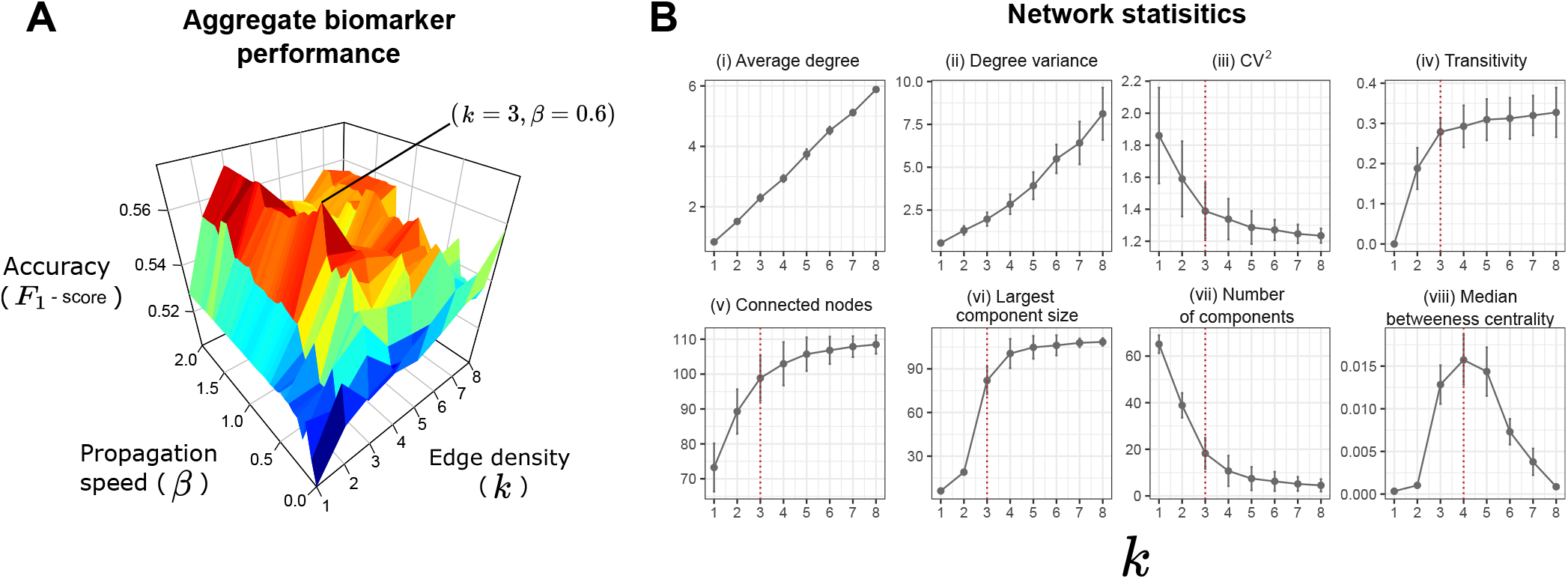
Optimisation of parameters on CPE_IMP_ and comparison to contact network topology. **(A)** Prediction accuracy (*F*_1_-score) as a function of propagation speed *β* and edge density *k*. Across all propagation speeds *β*, and edge density of *k* = 3 consistently maximises accuracy. Globally, accuracy peaks at a propagation speed of *β* = 0.6 and edge density *k* = 3. **(B)** Measures of network topology as a function of edge density: **(i)** average degree, **(ii)** degree variance, **(iii)** CV^2^, **(iv)** transitivity, **(v)** connected nodes, **(vi)** largest component size, **(vii)** number of components, **(viii)** and median betweenness centrality. For **(iii)** CV^2^, **(iv)** transitivity, **(v)** connected nodes, **(vii)** largest component size and **(viii)** number of components, we identified plateauing behaviour. Using an elbow detection algorithm^38^ we identified points of interest (the elbows) across the metrics exhibiting platues (**(iii), (iv), (v), (vi)**, and **(vii)**), highlighted by red vertical lines. In **(viii)** we identifyied spiking behaviour and instead highlighted the maximum in median betweenness centrality.

### Topological signals in the vicinity of optimal parameters

We further investigated the effect of edge density, which had a more profound effect on accuracy than propagation speed (cf. Figure 3A), on the contact structure as an indicator for underlying disease dynamics^43^ (Figure 3B). We quantified heterogeneity in the number of contacts of each individual by the squared coefficient of variation of the degree distribution (CV^2^)^44^, which can for example indicate the presence of ‘super-spreaders’ (a minority of individuals who infect many others^45^) and approximates how fast a disease will spread directly across a contact network^46^. The CV^2^ displayed an elbow at the optimal edge density *k* = 3, indicating that further increases in edge density do not largely change the pathogens spreading behaviour. Similarly, elbows of other topological network measures coincided with optimal edge density, namely, transitivity (a measure of how tightly connected nodes are, and a well-known proponent of network disease dynamics^47^), the number of connected nodes (patients with a connection to at least one other) and size of the largest component (both reflective of outbreak size), as well as the number of components (indicating the total number of distinct outbreak sub-clusters). The median betweenness centrality^48^, moreover, peaked close to the optimal density, suggesting that critical bridging nodes which connect communities^49^ are most identifiable across the contact network in this parameter region. Altogether, the sensitivity of network structure around optimal parameter values suggests that topological signals may indicate pathogen-specific parameters in an unsupervised setting when genomic ground-truth data is not available.

### Assessment of the contact network

To assess the trained model we compared its contact network *G*_*m*_ to two contact networks obtained via traditional methods: (i) the physical contact network (*G*_*p*_) with contacts between individuals who shared the same ward on the same day (see Methods: Standard contact model), and (ii) a ward-centric contact network *G*_*w*_ frequently used in the clinical setting (see Methods: Locationcentric contact model), where cases are linked if identified on the same ward within a +/- 7 day period^50^ (Figure 4A).

**Figure 4.**
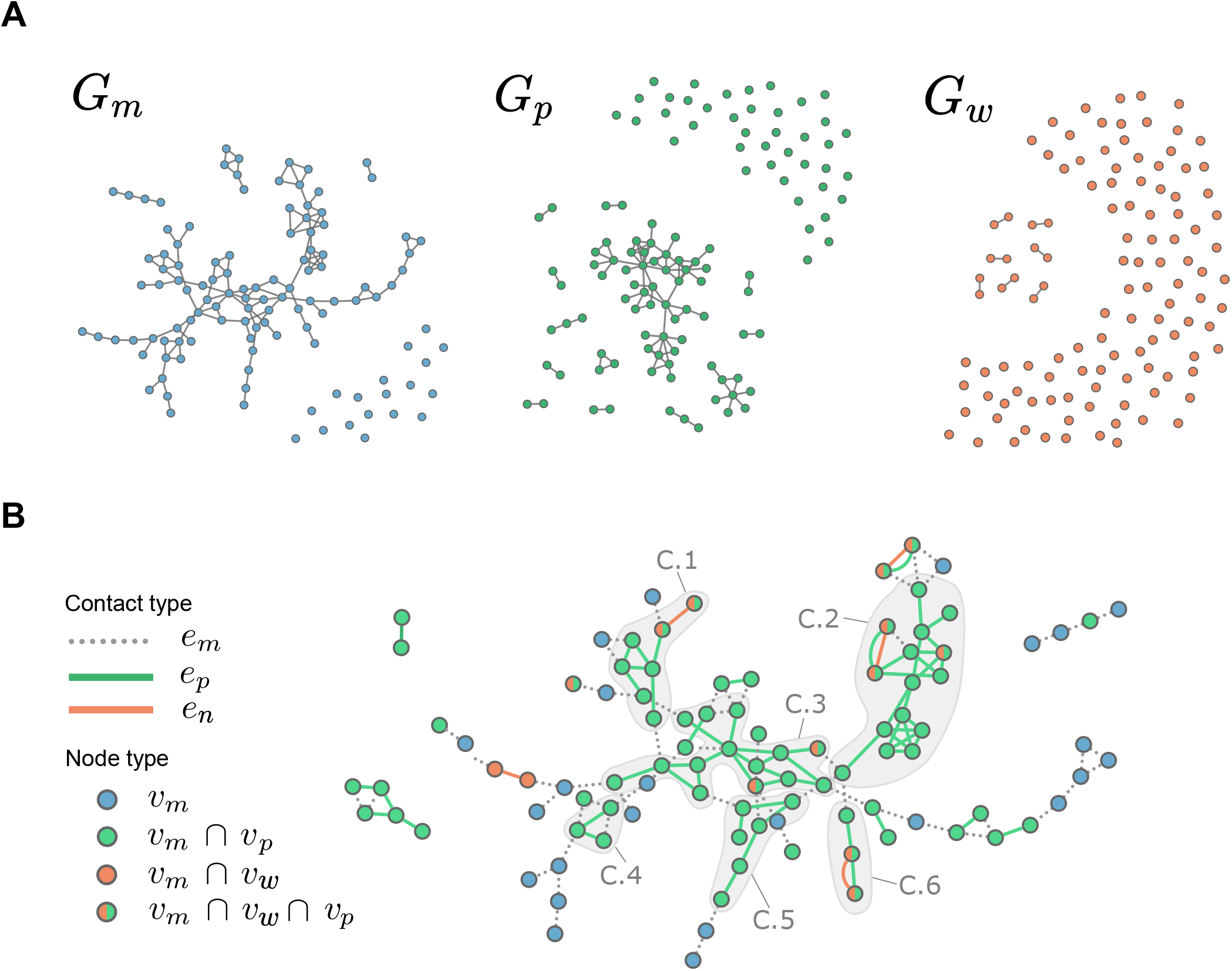
Comparison of contact networks. Contact networks for (1) *G*_*m*_ derived via StEP (which includes indirect contacts through background patient movement), (2) *G*_*p*_ via physical contact tracing, capturing contacts between patients shared a ward on the same day, and *G*_*w*_ constructed via ward-centric tracing (which defines contacts from cases identified on the same ward where within a +/- 7 day period). The contact networks identified by the three methods. **(B)** Overlap among network structures. *G*_*m*_ with edges and nodes coloured by their presence in *G*_*p*_ and/or *G*_*w*_. **(C**.**1) - (C**.**6)** highlight regions in the main component of *G*_*m*_ that contained 2 or more contacts from *G*_*p*_.

As StEP accounts for both direct and indirect contacts, unsurprisingly, it identified most contacts, followed by the physical contact network, and the ward-centric network identified only few contacts (*G*_*m*_: 123, *G*_*p*_: 88, *G*_*w*_: 7). The number of connected cases, consequently, varied substantially between networks (*G*_*m*_: 98/116, *G*_*p*_: 75/116, *G*_*w*_: 14/116). In particular, *G*_*m*_ revealed a large sub-network of highly connected cases. In contrast, *G*_*w*_ only identified few pairs of connected cases, which were predominately contained within larger connected components in *G*_*m*_ and *G*_*p*_ (Figure 4C). Overall, *G*_*m*_ contains only four disconnected components (compared to 13 in *G*_*p*_, and seven in *G*_*w*_), as well as the largest component comprised of 87 patients (compared to 42 in *G*_*p*_, and two in *G*_*w*_).

Although StEP can both introduce indirect and omit direct contacts, *G*_*m*_ largely contained structures of *G*_*p*_ and *G*_*w*_, including identified contacts (*G*_*p*_: 64/88, *G*_*w*_: 5/7) and connected patients (*G*_*p*_: 72/75, *G*_*w*_: 18/18, Figure S5), was also supported by an alignment of hierarchical clusterings of the weighted networks (Figure S4). Altogether, this means that StEP recovered the outbreak clusters identified via the traditional contact tracing methods and suggests that the extent of outbreaks may be largely underestimated when ignoring transmission via indirect contacts.

To test for alignment of the suggested outbreak structures with true transmission chains, we compared the contact networks to lineages obtained from sequencing of the IncHI2 plasmid, identified in 72 of the 85 sequenced isolates and found to be a driving force of the outbreak^35^(Figure S2). We used four community detection algorithms^51–54^ to partition patients into contact clusters, which we then compared to isolate clusters identified by the plasmid analysis (Table 1).

**Table 1.** Alignment of contact network to lineages of IncHI2 plasmids. Patient contact networks were clustered with four different community detection algorithms, and the resulting patient clusters were compared to plasmid lineages identified using genomics analysis. For each comparison, the Adjusted Rand Index (ARI)^55^ was computed, as well as a *p*-value which measures the significance of the alignment given a randomly assigned clustering of the same composition. Asterisks highlight significant *p*-values < 10^−2^.

| Clustering algorithm | Contact network | Adjusted Rand Index | $p$-value |
| --- | --- | --- | --- |
| Walktrap<sup>51</sup> | $G_m$ | 0.111 | $4.87 \times 10^{-8} *$ |
| | $G_p$ | 0.098 | $7.02 \times 10^{-7} *$ |
| | $G_w$ | -0.002 | $5.40 \times 10^{-1}$ |
| Edge betweenness<sup>52</sup> | $G_m$ | 0.076 | $1.05 \times 10^{-4} *$ |
| | $G_p$ | 0.079 | $5.24 \times 10^{-5} *$ |
| | $G_w$ | -0.002 | $5.30 \times 10^{-1}$ |
| Label propagation<sup>53</sup> | $G_m$ | 0.076 | $1.01 \times 10^{-5} *$ |
| | $G_p$ | 0.075 | $2.71 \times 10^{-4} *$ |
| | $G_w$ | -0.002 | $5.10 \times 10^{-1}$ |
| Fast-greedy<sup>54</sup> | $G_m$ | 0.048 | $7.05 \times 10^{-3} *$ |
| | $G_p$ | 0.057 | $3.80 \times 10^{-3} *$ |
| | $G_w$ | -0.002 | $5.14 \times 10^{-1}$ |

Due to low connectivity, *G*_*w*_ did not significantly align with plasmid lineages, regardless of the algorithm, while both *G*_*m*_ and *G*_*p*_ displayed better and significant alignment across all community detection algorithms (Table 1). Overall, *G*_*m*_ with its additional 23 connected patients over *G*_*p*_, achieved highest alignment to plasmid lineages (when using Walktrap^51^), supporting the extent of outbreak clusters via indirect contacts estimated by StEP. We therefore conclude that using a ward-centric contact tracing would wrongly suggest that outbreaks are a series of isolated transmission events. By considering patient trajectories, physical contact tracing improves on the ward-centric approach, however, best alignment with additional outbreak data is achieved without constraints on strictly observable physical contact.

### Case study 2: Applying the trained model to unseen outbreaks of CPE

We next applied the trained CPE_IMP_ model onto three unseen outbreaks of CPE carrying different resistance genes, namely (i) *bla*_OXA-48_ (CPE_OXA-48_), (ii) *bla*_NDM_ (CPE_NDM_), and (iii) *bla*_VIM_-family resistance genes (CPE_VIM_)(for an overview of these CPE’s we refer the reader to *Logan & Weinstein*^34^). As these outbreaks concern the same family of pathogen, *Enterobacteriaceae*, we assume that the epidemiological parameters optimised for CPE_IMP_ are applicable and transferable. We validate this hypothesis by (i) evaluating the alignment between the contact networks and bacterial species, and (ii) by analysing topological signals as previously in Case Study 1.

### Validation of StEP against bacterial species

Bacterial species shared by patients often used in the study’s hospital trust to quickly identify outbreaks of CPE between epidemiologically connected patients in the absence of genomic data. As species are a high-level summary and due to cross-species outbreaks driven by both species and plasmids, bacterial species, however, do not align perfectly with transmission events, rendering bacterial markers fuzzy indicators of transmission^56,57^. Nevertheless, for each CPE-type the StEP-derived contact networks *G*_*m*_ displayed better alignment with the distribution of bacterial species (and across all individual species labels Figure S9) than conventional the physical-contact graphs *G*_*p*_, again suggesting a substantial role of indirect contacts in transmission (Table 2).

**Table 2.**
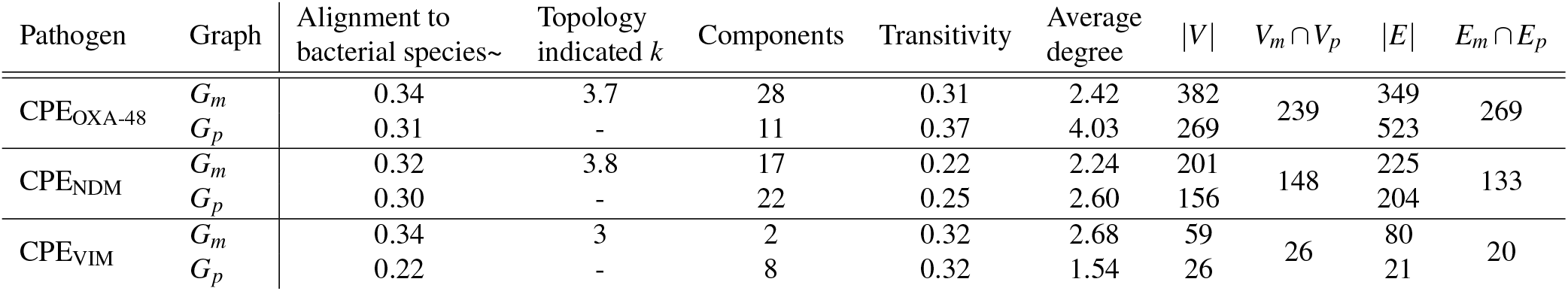
Summary statistics of CPE_OXA-48_, CPE_NDM_, and CPE_VIM_ contact network. For each pathogen, topological statistics are displayed for the StEP-derived contact network *G*_*m*_ and the physical contact network *G*_*p*_. In the first column, the average alignment with bacterial species is shown as computed using the label diffusion via GDR (determined using the *F*_1_-score from GDR, see Methods: GDR and Methods: Classification metric), where node labels are bacterial species (for breakdown by individual bacterial species see SI 3.2). A larger value indicates the contact network structure is more aligned to the distribution of bacterial species. The second column displays the ‘optimal’ edge density *k* found by considering each network measure (unsupervised parameter optimisation using elbow and peak detection averaged over topological measures, only applicable to the StEP-derived contact graph *G*_*m*_). The network statistics of the resulting contact graphs including number of components (not including isolated patients), transitivity, average degree, and number of nodes and edges are shown as well as the intersection of nodes and edges between both contact models.

| Pathogen | Graph | Alignment to bacterial species~ | Topology indicated $k$ | Components | Transitivity | Average degree | $\vert V\vert $ | $V_m \cap V_p$ | $\vert E\vert $ | $E_m \cap E_p$ |
| --- | --- | --- | --- | --- | --- | --- | --- | --- | --- | --- |
| CPE<sub>OXA-48</sub> | $G_m$ | 0.34 | 3.7 | 28 | 0.31 | 2.42 | 382 | 239 | 349 | 269 |
| | $G_p$ | 0.31 | - | 11 | 0.37 | 4.03 | 269 | | 523 | |
| CPE<sub>NDM</sub> | $G_m$ | 0.32 | 3.8 | 17 | 0.22 | 2.24 | 201 | 148 | 225 | 133 |
| | $G_p$ | 0.30 | - | 22 | 0.25 | 2.60 | 156 | | 204 | |
| CPE<sub>VIM</sub> | $G_m$ | 0.34 | 3 | 2 | 0.32 | 2.68 | 59 | 26 | 80 | 20 |
| | $G_p$ | 0.22 | - | 8 | 0.32 | 1.54 | 26 | | 21 | |

### Topologically indicated edge density

As in Case study 1, we investigated the topological sensitivity of the contact networks derived by StEP to edge density *k*. All CPE-types indeed displayed similar elbows and maxima in the network summary statistics to those observed for CPE_IMP_, i.e. around *k*≈ 3 (*k*∈ [3, 4] for all CPE-types, see Table 2 and Table S2), and for CPE_VIM_ all metrics suggested an optimal edge density of *k* = 3, same as for CPE_IMP_. This implies that network topologies are comparable to CPE_IMP_, in line with the outbreaks concerning the same family of pathogens with largely the same epidemiological characteristics.

### Comparison to traditional contact networks

Finally, we compared the contact networks derived from transferring the CPE_IMP_ StEP model (with propagation speed *β* = 0.6, and edge density *k* = 3) to the other CPE-types with contact networks derived from physical contact tracing (Figure S8). As for CPE_IMP_, StEP suggests a higher number of connected cases than the physical contact model across all other CPE-types (CPE_OXA-48_: 382 versus 269, CPE_NDM_: 201 versus 156, CPE_VIM_: 59 versus 26, Table 2). Connected cases in the physical contact network are largely contained within those identified by StEP (88.8%, 94.9%, and 100% for CPE_OXA-48_, CPE_NDM_, and CPE_VIM_ respectively), and despite differences in the number of connected patients, the hierarchical clusters of the StEP and physical contact networks aligned well (Figures S10-S12), suggesting that StEP recovered the core structures of a traditional contact model, while improving outbreak identification through inclusion of indirect contacts. Moreover, the increased scope of contact for each CPE type through StEP’s indirect links confirms undetected CPE carriage and transmission presents a significant burden amongst the hospital trust^58^.

### Case study 3: Exploring an outbreak of COVID-19 completely unsupervised

Our final Case Study examines 90 hospital patients who acquired COVID-19 during their hospital stay (patients identified in a previous study by *Price et al*.^59^). In the absence of additional genomic data, we previously transferred parameters from a pre-trained StEP model to unseen outbreak data of comparable pathogens. COVID-19, however, this presents a wholly different type of pathogen, that spreads with high dispersion via air droplets, compared to CPE, which typically spreads via shared touch surfaces^60,61^. We therefore learned pathogen-specific parameters by analysing the topological signals previously identified with CPE (see also Methods: Unsupervised learning of parameters).

As in Case study 1, edge density had a more pronounced effect than propagation speed on the topology of the contact network (Figure S14 & S14). Qualitatively, all metrics displayed comparable trends in edge density (Figure 5A), however, the locations of elbows and peaks across network metrics suggest a higher pathogen-specific edge density for COVID-19 (*k*∈ [4, 6], average *k* = 5). Lower edge densities resulted in disjoint clusters that only included highly weighted edges, i.e. the most likely transmission routes, which were embedded in further plausible transmission clusters in the contact networks resulting from higher edge densities (Figure 5B). Examining the contact network at the topologically indicated edge density, *k* = 5, we find a large component connecting 69/90 COVID-19 patients. As apposed to a location-centric method originally used in ref^59^), suggesting a far more linked and connected outbreak (SI XX). We also found strong alignment between the core contact structures of the StEP contact network *G*_*m*_ at *k* = 5 and the physical contact network *G*_*p*_ (Figure 5C). Even at granular scales, 70 out of 81 patients aligned within leaves of the hierarchical clustering, suggesting that physical contact structure was indeed preserved.

**Figure 5.**
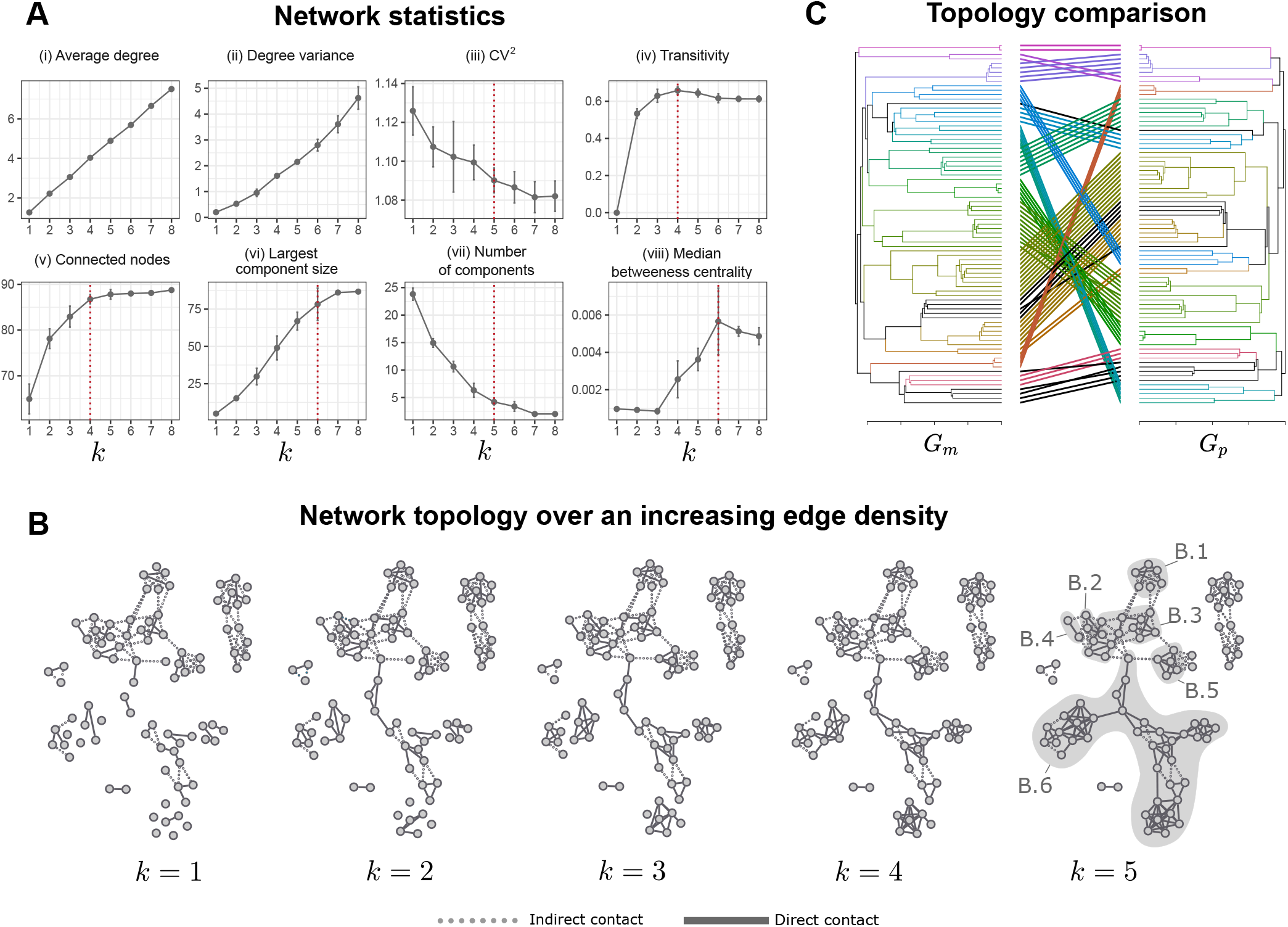
Summary of COVID-19 contact network from scans over StEP model parameters. Panel **A**: network statistics as a function of edge density: Average degree, degree variance, CV^2^, transitivity, number of nodes, largest component size, number of components, and the median betweenness centrality value. Panel **B**: contact network structure for edge density *k* = 1, …,5, (where propagation speed is fixed *β* = 0.4 where further increases had little effect on the inclusion of edges - Figure S14 & S14) with indirect contact and physical contact edges displayed as dotted and solid lines respectively. Sub panels **B1**.**1-B1**.**6** then highlight the physical contact structures within the largest component in the sparsity *k* = 5 contact network. Panel **C**: the topologically indicated optimal edge density *k* = 5 StEP contact network compared to the physical contact network in terms of hierarchical structure (recovered by the walktrap clustering algorithm^51^).

The elevated contact density identified by our analysis is consistent with COVID-19 presenting a highly transmissible pathogen^60^. With more than 75% of cases indicated to be connected, our analysis indicates a much larger extent of hospital transmission than suggested by traditional contact tracing, and thus crucially, highlights the importance of considering missing data and indirect contacts for infection control and prevention.

## Discussion

We introduced a graph-based model to recover pathogen-tailored contact networks amenable to tracing disease trans-mission. StEP integrates total movement into proximity measures to determine the likelihood of transmission between individuals who may or may not have had direct physical contact. It offers improved characterisation of transmission via more comprehensive contact networks. We used StEP to recover contacts among hospital patients in five disease outbreaks. Initially, we optimised StEP for an outbreak of one type of CPE, using genomic data available for a subset of cases^35^. We then deployed the optimised model on three unseen datasets of other CPE-types, and in the absence of genomic markers, validated its fitness with weaker transmission markers often used in clinical practice. Finally, we demonstrated how the model can be applied in an unsupervised setting to provide insights for outbreaks of novel pathogens, as presented by COVID-19 during the early stages of the pandemic. In all cases, StEP retrieved more extensive contact networks that preserved core physical contact structures and, where available, better aligned with independent outbreak data, altogether implying a larger extent of hospital transmission than identified by traditional methods.

StEP presents promising advances for contact network epidemiology, where proximity sensors have become a gold-standard to collect detailed contact data and understand infectious disease transmission. ^62–66^. The underlying assumption is that proximity increases the likelihood of disease transmission, and we find support for this assumption through the alignment of genomic variation with spatial-temporal proximity (SI 2.3). Proximity-based methods can, however, miss certain transmission events facilitated by unobserved individuals, for example, healthcare staff or visitors, or indirectly, for example, via contaminated instruments and surfaces. StEP addresses missed contacts by considering total movement as the key vector for disease transmission, an assumption that has proven effective for various epidemiological scenarios^22, 23, 25, 26, 28–31^.

The model can be trained to pathogen-specific transmission features, and the learned parameters can give insights into epidemiological differences between pathogens such as the level of contact (space time proximity) leading to transmission^64^. Once tuned for a general class of pathogens, StEP can moreover be deployed in real-time and unsupervised, without the need for additional genomic sequencing, which offers a major advantage over alternative reconstruction models that integrate both genomic and epidemiological data^67,68^. In addition, the graph-based framework is robust to missing data, as often the case in real-world scenarios with only partially complete datasets.

Our analysis suggests that pathogen-tailored contact networks feature topological signatures associated with plausible parameter regions. Across all networks and epidemiological parameters, we observed some highly connected individuals, likely the result of heterogeneity in infectiousness^45^. For specific parameter values, moreover, we observed bridging individuals, highlighted by their pronounced betweeness centralities^49^. Importantly, optimal parameters coincided with topological changes across a number of network metrics related to disease dynamics. For COVID-19, the topological signals correctly identified an elevated contact density and thus higher transmissibility. Such information may guide investigations of other newly emergent pathogens, when data is typically scarce.

In this work, we considered static background movement, however, in practice, background movement is likely non-static, for example, due to time-varying fluctuations in human mobility^69^, or more disruptive changes, such as the ongoing COVID-19 pandemic^23,70^. One way to incorporate such temporal information is by replacing the static background movement in StEP for a temporal network. In this setting, the shortest path in the proximity kernel becomes the shortest time-respective pathway^71^ between two locations. The undirected networks recovered by StEP contain contacts that likely led to transmission, but not its direction^1^. Possible future extensions of StEP may account for directed edges to recover transmission trees and provide further granularity to inform outbreak analyses. Furthermore, application of StEP to broader classes of pathogens and validation with WGS would further demonstrate the model’s generalisability.

Overall, StEP tackles several challenges associated with contact tracing, and our analysis highlights the importance of indirect (and/or unobserved) contacts in disease transmission. Our approach takes both direct and indirect contacts into account and thus provides an extensive characterisation of disease outbreaks. Its flexible use of heterogeneous data can significantly enhance insights to inform interventions for infection control and prevention.

## Methods

### Movement data acquisition

Our analysis is based on anonymised electronic health records of patients from a large 1000-bed Trust of teaching hospitals in London. Total hospital movement patterns were constructed from all hospital patients’ (1862) inter-ward movements (not just those with an infection) over a month of regular hospital activity (see SI 1 - Background hospital movement network). For Case study 1, which looked at CPE_IMP_, the known CPE_IMP_-carrying patients pathways (116) were obtained during the original study duration (March 2016 to December 2019) in *Boonyasiri et al*.^35^. Case study 2 looking at CPE_OXA-48_, CPE_NDM_, CPE_VIM_, looked at patient carriage (for each type) between August 2018 and August 2020. For greater overview on background CPE during the study duration, we direct the reader to a local study in the same hospital trust by *Otter et al*.^58^, or to a national overview by *Public Health England*^41^. Case study 3, which looked at COVID-19 in the same hospital teaching trust, studied the first wave of COVID-19 in the UK between March and April in 2020 (detailed in *Price et al*.^59^).

### Genomics analysis of IncHI2 plasmids

WGS reads of the 85 CPE_IMP_ isolates were processed and analysed for plasmid detection, phylogenetic reconstruction, and plasmid-lineage identification, as we have described in another study^35^. Briefly, using IQ-Tree^72^, a maximum-likelihood phylogenetic tree of IncHI2 plasmids were reconstructed from whole-genome alignment of the reads against a reference plasmid genome pKA_P10 (RefSeq accession: NZ_CP044215.1)^73^. Then the tree was corrected for recombination using ClonalFrameML and eventually was rooted using BactDating^74,75^. Plasmid lineages were identified from the rooted tree based on its topology and bootstrap values (≥65) in the output maximum-likelihood tree of IQ-Tree (Figure S2). Database ResFinder (updated on 28 Oct 2020) and software ARIBA v2.14.6 were used for detecting acquired antimicrobial resistance genes from the reads of 72 isolates (out of the 85 isolates) carrying IncHI2 plasmids^76,77^. Alleles of identified acquired antimicrobial resistance genes were determined and labelled using GeneMates helper script PAMmaker v0.0.5, which also created a presence-absence matrix of these alleles across the 72 isolates^78^.

### Biomarker construction

To track the spread of CPE_IMP_, 85 available CPE isolates from 116 confirmed CPE_IMP_-carrying patients were sampled for WGS and the detection of acquired resistance genes. In total, 106 alleles were identified, forming 181 three-allele combinations that we considered as biomarkers (Table S1). For an isolate to have a particular biomarker, all three alleles must have been found present together in their genomic analysis.

### Effective distance and probable paths of disease

Routes of disease spread are often dominated by a set of most probable trajectories in terms of *effective distance*^36^. To derive such effective distance trajectories, we firstly construct a transition matrix *P* which captures background system movement. In *P*, elements *P*_*i j*_ confer to the fraction of departing individuals leaving node *v*_*i*_ and arriving at node *v* _*j*_, such that 1 *< P*_*i j*_ ≤ 1. Given *P* effective distance *d*_*i*_ *j* is,

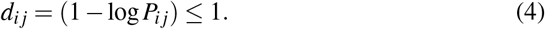

In *d*_*i*_ *j* a low proportion of movement from *v*_*i*_ →*v* _*j*_ corresponds to a large effective distance. Explained in greater detail in ref^36^, the logarithm captures additive effective distances along multi-step pathways. The most probable trajectories of disease spread from *v*_*i*_ → *v*_*j*_ is given by the directed path *δ*_*i j*_ ={*v*_*i*_ →, …, →*v* _*j*_}with the smallest total effective distance *λ* (Γ):

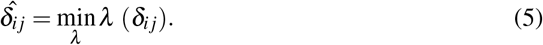

Typically, 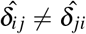, since movement patterns vary depending localities of the underling system. Also note, for paths a shortest path 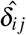 may not exist (when no path is observed).

### Continuous *k* -nearest neighbours

CkNN^39^ is a density aware neighbourhood joining methods. Whilst CkNN is predominantly developed for graph construction, we have adapted it here for filtering graph edges since it provides a geometrically consistent manifold representation. In this paper we implement CkNN to construct a contact network in two steps: Firstly, we define pairwise distance between any two nodes *m* and *n* as *d*(*n, m*). Secondly, we use *d*(*n, m*) in CKNN to define the CKNN graph as follows:

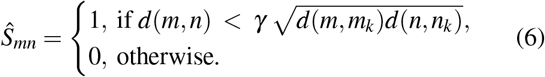

Here *n*_*k*_ and *m*_*k*_ represent the *k*-th nearest neighbours for samples *n* and *m* respectively. Importantly, the two parameters *k* (ranging from 1 to *N*-1), and *γ* regulate the CKNN graph structure. Parameter *k* regulates the *k* nearest nearest neighbours to consider and hence network sparsity, whereas *γ* is a positive parameter regulating local point density. In this work we focused primarily on *k* by fixing *γ* = 1.

### Semi-supervised classification on graph using diffusion dynamics

Graph Diffusion Reclassification (GDR) leverages explicit diffusive dynamics through of the graph through its Laplacian^42^. GDR uses continuous-time graph diffusion of each class label for the training nodes as a means to classify the set of evaluation nodes. Through searching the time evolution of node dynamics we can identify the maximum probability of class assignment which forms the basis to classify the test nodes. Here, only the node classes are defined and node features are not present (node features can be used to define a prior on the class probability, however, without node features the probability is equal for each class).

Starting with a prior assignment matrix *ℋ*, which here is taken to be a flat distribution of equal probability of being in each class, a node is classified according to the maximum probability (here described as node overshoots) of any of the *c* classes. It is easy to see that the values of all the node overshoots are captured compactly in matrix form as

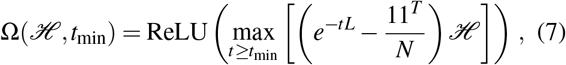

and the node reclassification is given from the maximum over-shoot across classes

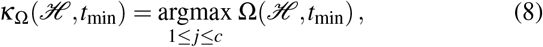

where the argmax is the standard row-wise operator that finds the maximum across classes, and we define argmax(0) = so that the indicator vector 𝟙_0_(*κ*_Ω_) marks the set of non-overshooting nodes. The GDR code is available at github.com/barahona-research-group/GDR).

### Classification metric

The classification accuracy from GDR is then reported as the *F*_1_-score across all biomarkers and averaged over predictions for five test sets of Monte-Carlo Cross Validation^79^:

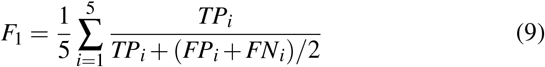

where *TP*_*i*_, *FP*_*i*_, and *FN*_*i*_ represent true-positive, false-positive, and false-negative rates, respectively, of the *i*-th test set, when compared presence-absence of biomarkers predicted from network topology to their actual distributions amongst CPE_IMP_ isolates.

### Learning disease-specific StEP parameters

The StEP model features two key parameters, namely the propagation speed *β* and edge density *k*. These parameters reflect the transmission dynamics of the pathogen under consideration. We present two approaches to identify StEP parameters: firstly, a supervised approach based on partially labelled data that aligns the contact network to transmission markers using a predictive framework, representing the more sensitive means to obtain optimal parameters; and secondly, an unsupervised approach based on the structural properties of the StEP contact network, which is less sensitive, but aligns the network to edge density.

### Semi-supervised learning of parameters

To learn parameters, we use a semi-supervised node classification framework, Graph Diffusion Reclassification (GDR)^42^ to obtain a network alignment to transmission by predicting node labels (see Methods: GDR). Whilst in this study we use genetic biomarkers as node labels, and as a means to trace pathogen transmission, the StEP methodology is general and amenable to alternative markers that follow transmission. Because transmission between two individuals is likely to have occurred if their pathogens contained similar sets of biomarkers markers, this predictive approach can identify a contact network best aligned to transmission. Importantly, this semi-supervised framework is robust to missing data which is standard for real-world epidemiological investigations under limited resources. For the final classification accuracy of GDR we reported the *F*_1_-score across all biomarkers, averaged over predictions across five test sets of Monte-Carlo Cross Validation^79^ (see Methods: Classification metric).

### Unsupervised learning of parameters

In many instances, ground truth labels are not directly accessible, precluding the use of supervised methods. Unsupervised techniques exploit the intrinsic structure present in the input data to identify model parameters^38^. In this study, as well as directly learning parameters from labelled data, we demonstrate how network topological metrics, namely plateaus and spikes in network metrics, provide signal aligning to tuned parameters and validated parameters. These signals reflect pathogen-specific contact structure, namely reflecting direct infectiousness, and offer an unsupervised means to explore contact in the early stages of new disease outbreaks.

### Alternative contact network construction

#### Standard contact model

Standard approaches to contact recovery look for observable contact when two individuals are in the same location at the same time to established links^13^. This means contact is established between any two individuals if their trajectories *T*_*n*_ and *T*_*m*_, if any of their elements (the location-timing *l*_*i*_ = (*v*_*i*_, *t*_*i*_), see Equation 1) intersect |*T*_*n*_ ∩ *T*_*m*_| ≠ ø. Thus, the standard contact network for a set of trajectories can be represented by *N* ×*N* adjacency matrix *S* with elements *S*_*nm*_ ≠ 0 if two individuals were in direct contact. Moreover, non-zero elements of *S* are weighted by contact duration (cardiniality of the set intersection),

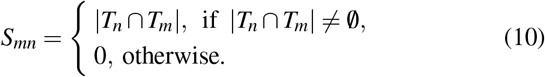

#### Location-centric contact model

In operational settings, standard contact definition can be challenging to fully implement in hospital due to the large number of linked patients generated by contact with identified carries. Often it is more feasible to restrict the definition of contact, and link cases in space and time bases on where they have been identified^50^.For some pathogens, these points my fall in locations and time where they were screened, or where they became symptomatic time as was used for COVID-19 previously in^59^. Namely, a location-centric approach is taken, which establishes links between individual if they had been identified close across time *t* in the same location *v*_*i*_.

For each individual a single location-timing tuple, *p*_*i*_ = (*v*_*i*_, *t*_*i*_), records where (*v*_*i*_) and when (*t*_*i*_) they were identified as a positive carrier. Formally, location-centric contact is established between two individuals if they have been in the same location within Δ*t* amount of time from when either was identified as positive. Thus the adjacency matrix *S* between *N* individuals based on location-centric contact is defined as:

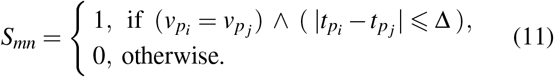

### COVID-19 trajectory pre-processing

An importance different between COVID-19 and CPE is the shorter incubation time of COVID-19. Therefore, when constructing the patient trajectories for COVID-19, we only considered movement 14-days before diagnosis^80^ (this is a choice that would also be necessary for the traditional contact model, not a limitation of StEP).

## Data Availability

The patient pathway datasets generated and analysed specifically for the study are not publicly available to protect anonymity of included hospital patients. However, for datasets incorporated from existing studies, we refer readers to \textit{Boonyasiri et al.}~\cite{IMP2021} for Case study 1 data on CPE\textsubscript{IMP}, and \textit{Price et al.}\cite{Price_Mookerjee2020} for Case study 3 data on hospital acquired COVID-19.

https://github.com/ashm97/StEP

## Additional information

### Data collection and ethics

Study ethics were granted and approved through Imperial College London NHS Trust service evaluations (Ref:386,379,473). All patient pathway data was collected from the central business intelligence system and fully pseudanonymised before analysis, in accordance with ethics 15_LO_0746.

### Availability of data and materials

The patient pathway datasets generated and analysed specifically for the study are not publicly available to protect anonymity of included hospital patients. However, for datasets incorporated from existing studies, we refer readers to *Boonyasiri et al*.^35^ for Case study 1 data on CPE_IMP_, and *Price et al*.^59^ for Case study 3 data on hospital acquired COVID-19.

The repository for StEP can be found at github.com/ashm97/StEP and GDR at github.com/barahona-research-group/GDR.

### Author contributions statement

AM performed main computational analyses. RP and AM implemented semi-supervised model optimisation. YW conducted genomics analyses. AM wrote computer code and YW made the StEP R package available. AM, RP, and MB contributed to StEP methodological development. YW, SM, EJ, JP, AH, and FD provided interpretation of data and results. AM, RP, YW, AYW, and MB wrote the manuscript. AYW, AH, and MB provided project supervision. All authors have read and approved this publication.

## Acknowledgements

We wish to thank Eleonora Dyakova for help with accessing data.

## Supplementary information

## 1 StEP model

### 1.1 Background hospital movement network

The total mobility network incorporated by StEP in this study (for all case studies) is constructed from 4902 inter-ward transfers of 1862 hospital patients. This data set constitute a large sample of movement patterns from 1862 from 2018 to 2020 and encompassed 129 hospital wards across 5 geographically separate sites. Naturally, our mobility data is represented by of directed graph *G* = {*V, E*}, in which nodes *V* = *v*_*i*_ represent hospital wards and edges *E* = *v*_*i j*_ the directed movement of patient between wards *v*_*i*_ →*v* _*j*_. Introducing weighting on edges, as *w*_*i j*_ then allows us to capture total volume of patients moving from *v*_*i*_ →*v* _*j*_ in the sampled data. The resultant *G* contained 1461 total directed edges (Figure S1). On average nodes in *G* had an in-degree and out-degree of 11. Previous analysis of the same hospital by *Myall et al*.^1^ found communities conferring to both different hospital departments and physical structures (geographical sites and different buildings). This correspondence to known structures suggests an ability graph captures hospital movement patterns, as well as some amount of staff movement given the alignment to different departments.

**Figure S1.**
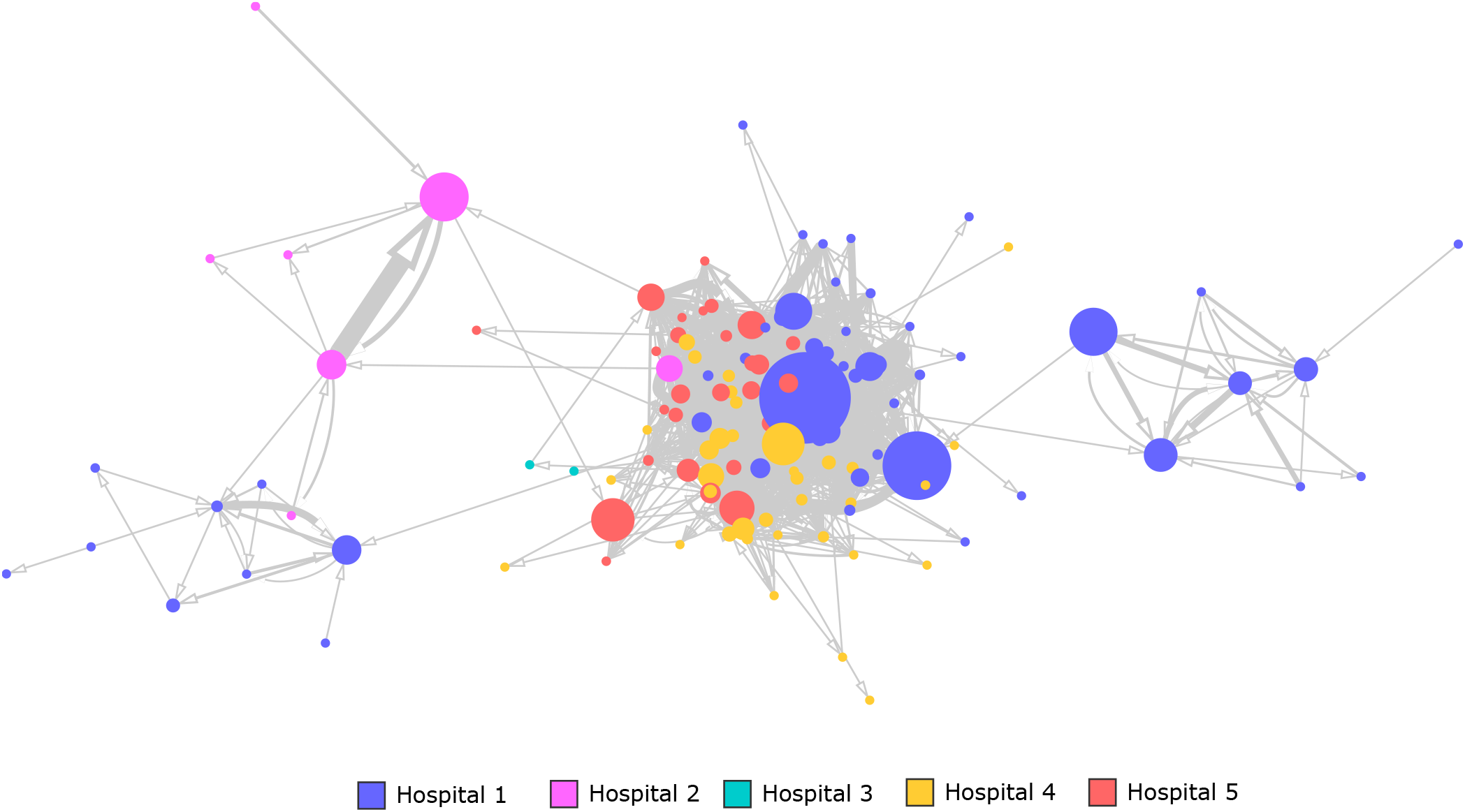
Directed hospital mobility network. The hospital network is constructed from patient movements from wards *v*_*i*_ →*v* _*j*_. Nodes are scaled by network betweenness centrality, and edge widths are sized according to total number of patient transfers. Node colouring refers to which of the five hospital sites the ward belonged too.

## 2 Case study 1

### 2.1 Biomarkers

CPE_IMP_ biomarkers were recovered from Whole Genome Sequencing of pathogen isolates. For each sample the presenceabsence of 106 alleles of acquired antimicrobial resistance genes were recorded in a feature matrix^2^. From this allele feature matrix, we engineered a set of 181 biomarkers, based on the co-occurrence of three different alleles in a CPE_IMP_ isolate (Table S1). Whilst a possible 5831820 combinations of three biomarkers existed, we restricted our set of interest such that the class distribution was largely equal and was no worse than 80-20 (a total present class count between 15 and 57).

**Table S1.** Biomarker table. Each biomarker represents co-occurrence of three different alleles of acquired antimicrobial resistance (AMR) genes in a CPE_IMP_ isolate. Each suffix ‘var’ of allele labels was assigned by PAMmaker to represent a specific variant of a reference allele in the ResFinder database^3,4^. In GDR’s label diffusion framework, each biomarker is diffused over the network structure, giving a classification accruacy (*F*_1_-score) which is averaged over predictions across five test sets of Monte-Carlo Cross Validation^5^.

| Biomarker | Allele 1 | Allele 2 | Allele 3 | mean | med | sd | count |
| --- | --- | --- | --- | --- | --- | --- | --- |
| biomarker_1 | blaSHV-12 | blaTEM-1B | sul1_2 | 0.74 | 0.77 | 0.12 | 29 |
| biomarker_2 | aph(3'')-Ib_5.var1 | aph(6)-Id_1 | blaSHV-12 | 0.73 | 0.91 | 0.24 | 28 |
| biomarker_3 | aph(3'')-Ib_5.var1 | blaSHV-12 | blaTEM-1B | 0.73 | 0.91 | 0.24 | 28 |
| biomarker_4 | aph(3'')-Ib_5.var1 | blaSHV-12 | mcr-9 | 0.73 | 0.91 | 0.24 | 28 |
| biomarker_5 | aph(3'')-Ib_5.var1 | blaSHV-12 | sul1_2 | 0.73 | 0.91 | 0.24 | 28 |
| biomarker_6 | aph(6)-Id_1 | blaSHV-12 | blaTEM-1B | 0.73 | 0.91 | 0.24 | 28 |
| biomarker_7 | aph(6)-Id_1 | blaSHV-12 | mcr-9 | 0.73 | 0.91 | 0.24 | 28 |
| biomarker_8 | aph(6)-Id_1 | blaSHV-12 | sul1_2 | 0.73 | 0.91 | 0.24 | 28 |
| biomarker_9 | blaSHV-12 | blaTEM-1B | mcr-9 | 0.73 | 0.91 | 0.24 | 28 |
| biomarker_10 | blaSHV-12 | mcr-9 | sul1_2 | 0.73 | 0.91 | 0.24 | 28 |
| biomarker_11 | blaTEM-1B | mcr-9 | qnrA1.var1 | 0.67 | 0.64 | 0.08 | 54 |
| biomarker_12 | aac(6')-IIc | catA2.var1 | dfrA19 | 0.67 | 0.66 | 0.06 | 34 |
| biomarker_13 | aac(6')-IIc | catA2.var1 | mcr-9 | 0.65 | 0.64 | 0.03 | 34 |
| biomarker_14 | aac(6')-IIc | catA2.var1 | qnrA1.var1 | 0.65 | 0.65 | 0.04 | 35 |
| biomarker_15 | aac(6')-Ib3.var1 | blaTEM-1B | catA2.var1 | 0.64 | 0.63 | 0.05 | 47 |
| biomarker_16 | aac(6')-IIc | blaIMP-70.var1 | catA2.var1 | 0.64 | 0.63 | 0.04 | 36 |
| biomarker_17 | aac(6')-IIc | blaTEM-1B | catA2.var1 | 0.64 | 0.63 | 0.04 | 36 |
| biomarker_18 | aac(6')-IIc | catA2.var1 | sul1_2 | 0.64 | 0.63 | 0.04 | 36 |
| biomarker_19 | aph(6)-Id_1 | blaIMP-70.var1 | catA2.var1 | 0.64 | 0.65 | 0.08 | 48 |
| biomarker_20 | aph(6)-Id_1 | blaTEM-1B | catA2.var1 | 0.64 | 0.65 | 0.08 | 48 |
| biomarker_21 | aph(3'')-Ib_5.var1 | mcr-9 | qnrA1.var1 | 0.64 | 0.66 | 0.1 | 55 |
| biomarker_22 | blaIMP-70.var1 | mcr-9 | qnrA1.var1 | 0.64 | 0.66 | 0.1 | 55 |
| biomarker_23 | dfrA19 | mcr-9 | qnrA1.var1 | 0.64 | 0.66 | 0.1 | 55 |
| biomarker_24 | mcr-9 | qnrA1.var1 | sul1_2 | 0.64 | 0.66 | 0.1 | 55 |
| biomarker_25 | aac(6')-IIc | aph(3'')-Ib_5.var1 | catA2.var1 | 0.63 | 0.63 | 0.05 | 35 |
| biomarker_26 | aac(6')-IIc | aph(6)-Id_1 | catA2.var1 | 0.63 | 0.63 | 0.05 | 35 |
| biomarker_27 | blaIMP-70.var1 | blaTEM-1B | catA2.var1 | 0.63 | 0.63 | 0.08 | 50 |
| biomarker_28 | catA2.var1 | mcr-9 | qnrA1.var1 | 0.63 | 0.64 | 0.04 | 47 |
| biomarker_29 | aph(3'')-Ib_5.var1 | catA2.var1 | qnrA1.var1 | 0.62 | 0.59 | 0.06 | 48 |
| biomarker_30 | catA2.var1 | dfrA19 | qnrA1.var1 | 0.62 | 0.59 | 0.06 | 48 |
| biomarker_31 | catA2.var1 | dfrA19 | mcr-9 | 0.62 | 0.63 | 0.08 | 49 |
| biomarker_32 | blaTEM-1B | catA2.var1 | qnrA1.var1 | 0.62 | 0.63 | 0.08 | 48 |
| biomarker_33 | aph(6)-Id_1 | catA2.var1 | qnrA1.var1 | 0.62 | 0.64 | 0.06 | 46 |
| biomarker_34 | aac(6')-Ib3.var1 | aac(6')-IIc | catA2.var1 | 0.61 | 0.59 | 0.07 | 34 |
| biomarker_35 | blaIMP-70.var1 | catA2.var1 | mcr-9 | 0.61 | 0.59 | 0.1 | 49 |
| biomarker_36 | aph(6)-Id_1 | catA2.var1 | mcr-9 | 0.61 | 0.59 | 0.06 | 48 |
| biomarker_37 | aac(6')-IIc | aph(3'')-Ib_5.var1 | mcr-9 | 0.61 | 0.61 | 0.01 | 38 |
| biomarker_38 | aac(6')-IIc | aph(6)-Id_1 | mcr-9 | 0.61 | 0.61 | 0.01 | 38 |
| biomarker_39 | aac(6')-IIc | blaIMP-70.var1 | mcr-9 | 0.61 | 0.61 | 0.01 | 38 |
| biomarker_40 | aac(6')-IIc | blaTEM-1B | mcr-9 | 0.61 | 0.61 | 0.01 | 38 |
| biomarker_41 | aac(6')-IIc | mcr-9 | sul1_2 | 0.61 | 0.61 | 0.01 | 38 |
| biomarker_42 | aac(6')-IIc | blaIMP-70.var1 | blaTEM-1B | 0.61 | 0.63 | 0.06 | 40 |
| biomarker_43 | aac(6')-IIc | blaIMP-70.var1 | sul1_2 | 0.61 | 0.63 | 0.06 | 40 |
| biomarker_44 | aac(6')-IIc | blaTEM-1B | sul1_2 | 0.61 | 0.63 | 0.06 | 40 |
| biomarker_45 | aac(6')-Ib3.var1 | blaIMP-70.var1 | catA2.var1 | 0.6 | 0.56 | 0.1 | 48 |
| biomarker_46 | aac(6')-Ib3.var1 | catA2.var1 | sul1_2 | 0.6 | 0.56 | 0.1 | 48 |
| biomarker_47 | aac(6')-IIc | dfrA19 | mcr-9 | 0.6 | 0.59 | 0.06 | 37 |
| biomarker_48 | aac(6')-IIc | mcr-9 | qnrA1.var1 | 0.6 | 0.59 | 0.06 | 37 |
| biomarker_49 | blaTEM-1B | catA2.var1 | mcr-9 | 0.6 | 0.59 | 0.1 | 49 |
| biomarker_50 | aph(3'')-Ib_5.var1 | blaIMP-70.var1 | qnrA1.var1 | 0.6 | 0.6 | 0.03 | 56 |
| biomarker_51 | aph(3'')-Ib_5.var1 | dfrA19 | qnrA1.var1 | 0.6 | 0.6 | 0.03 | 56 |
| biomarker_52 | aph(3'')-Ib_5.var1 | qnrA1.var1 | sul1_2 | 0.6 | 0.6 | 0.03 | 56 |
| biomarker_53 | blaIMP-70.var1 | dfrA19 | qnrA1.var1 | 0.6 | 0.6 | 0.03 | 56 |
| biomarker_54 | dfrA19 | qnrA1.var1 | sul1_2 | 0.6 | 0.6 | 0.03 | 56 |
| biomarker_55 | aph(3'')-Ib_5.var1 | catA2.var1 | sul1_2 | 0.6 | 0.61 | 0.08 | 51 |
| biomarker_56 | blaIMP-70.var1 | catA2.var1 | sul1_2 | 0.6 | 0.62 | 0.1 | 51 |
| biomarker_57 | blaTEM-1B | catA2.var1 | sul1_2 | 0.6 | 0.62 | 0.1 | 51 |
| biomarker_58 | aph(3'')-Ib_5.var1 | catA2.var1 | mcr-9 | 0.6 | 0.62 | 0.07 | 50 |
| biomarker_59 | catA2.var1 | mcr-9 | sul1_2 | 0.6 | 0.62 | 0.07 | 50 |
| biomarker_60 | blaTEM-1B | catA2.var1 | dfrA19 | 0.59 | 0.54 | 0.09 | 49 |
| biomarker_61 | aph(3'')-Ib_5.var1 | catA2.var1 | dfrA19 | 0.59 | 0.58 | 0.06 | 50 |
| biomarker_62 | catA2.var1 | dfrA19 | sul1_2 | 0.59 | 0.58 | 0.06 | 50 |
| biomarker_63 | aac(6')-Ib3.var1 | aph(3'')-Ib_5.var1 | catA2.var1 | 0.59 | 0.58 | 0.04 | 47 |
| biomarker_64 | aph(3'')-Ib_5.var1 | blaTEM-1B | catA2.var1 | 0.59 | 0.6 | 0.09 | 50 |
| biomarker_65 | aac(6')-Ib3.var1 | blaIMP-70.var1 | qnrA1.var1 | 0.59 | 0.64 | 0.12 | 55 |
| biomarker_66 | aac(6')-Ib3.var1 | qnrA1.var1 | sul1_2 | 0.59 | 0.64 | 0.12 | 55 |
| biomarker_67 | blaIMP-70.var1 | catA2.var1 | dfrA19 | 0.58 | 0.54 | 0.07 | 49 |
| biomarker_68 | aph(6)-Id_1 | blaIMP-70.var1 | mcr-9 | 0.58 | 0.56 | 0.09 | 54 |
| biomarker_69 | aac(6')-Ib3.var1 | aac(6')-IIc | blaIMP-70.var1 | 0.58 | 0.57 | 0.08 | 38 |
| biomarker_70 | aac(6')-Ib3.var1 | aac(6')-IIc | blaTEM-1B | 0.58 | 0.57 | 0.08 | 38 |
| biomarker_71 | aac(6')-Ib3.var1 | aac(6')-IIc | sul1_2 | 0.58 | 0.57 | 0.08 | 38 |
| biomarker_72 | aac(6')-Ib3.var1 | catA2.var1 | mcr-9 | 0.58 | 0.57 | 0.06 | 46 |
| biomarker_73 | aph(3'')-Ib_5.var1 | aph(6)-Id_1 | catA2.var1 | 0.58 | 0.58 | 0.08 | 49 |
| biomarker_74 | aph(6)-Id_1 | catA2.var1 | sul1_2 | 0.58 | 0.58 | 0.08 | 49 |
| biomarker_75 | aph(6)-Id_1 | blaIMP-70.var1 | blaTEM-1B | 0.58 | 0.59 | 0.05 | 54 |
| biomarker_76 | aac(6')-Ib3.var1 | catA2.var1 | qnrA1.var1 | 0.57 | 0.54 | 0.05 | 46 |
| biomarker_77 | aac(6')-Ib3.var1 | aac(6')-IIc | aph(3'')-Ib_5.var1 | 0.57 | 0.56 | 0.06 | 37 |
| biomarker_78 | aac(6')-Ib3.var1 | aac(6')-IIc | aph(6)-Id_1 | 0.57 | 0.56 | 0.06 | 37 |
| biomarker_79 | aph(3'')-Ib_5.var1 | blaTEM-1B | qnrA1.var1 | 0.57 | 0.56 | 0.13 | 55 |
| biomarker_80 | blaTEM-1B | dfrA19 | qnrA1.var1 | 0.57 | 0.56 | 0.13 | 55 |
| biomarker_81 | aph(3'')-Ib_5.var1 | aph(6)-Id_1 | qnrA1.var1 | 0.57 | 0.56 | 0.06 | 53 |
| biomarker_82 | aph(6)-Id_1 | blaIMP-70.var1 | qnrA1.var1 | 0.57 | 0.56 | 0.06 | 53 |
| biomarker_83 | aph(6)-Id_1 | dfrA19 | qnrA1.var1 | 0.57 | 0.56 | 0.06 | 53 |
| biomarker_84 | aph(6)-Id_1 | qnrA1.var1 | sul1_2 | 0.57 | 0.56 | 0.06 | 53 |
| biomarker_85 | aac(6')-IIc | blaIMP-70.var1 | qnrA1.var1 | 0.57 | 0.57 | 0.08 | 39 |
| biomarker_86 | aac(6')-IIc | blaTEM-1B | qnrA1.var1 | 0.57 | 0.57 | 0.08 | 39 |
| biomarker_87 | aac(6')-IIc | qnrA1.var1 | sul1_2 | 0.57 | 0.57 | 0.08 | 39 |
| biomarker_88 | aph(3'')-Ib_5.var1 | blaIMP-70.var1 | catA2.var1 | 0.57 | 0.6 | 0.06 | 50 |
| biomarker_89 | aac(6')-Ib3.var1 | aph(6)-Id_1 | mcr-9 | 0.57 | 0.6 | 0.07 | 55 |
| biomarker_90 | aac(6')-IIc | aph(3'')-Ib_5.var1 | dfrA19 | 0.56 | 0.54 | 0.1 | 38 |
| biomarker_91 | aac(6')-IIc | aph(3'')-Ib_5.var1 | qnrA1.var1 | 0.56 | 0.54 | 0.1 | 38 |
| biomarker_92 | aac(6')-IIc | aph(6)-Id_1 | dfrA19 | 0.56 | 0.54 | 0.1 | 38 |
| biomarker_93 | aac(6')-IIc | aph(6)-Id_1 | qnrA1.var1 | 0.56 | 0.54 | 0.1 | 38 |
| biomarker_94 | aac(6')-IIc | blaIMP-70.var1 | dfrA19 | 0.56 | 0.54 | 0.1 | 38 |
| biomarker_95 | aac(6')-IIc | blaTEM-1B | dfrA19 | 0.56 | 0.54 | 0.1 | 38 |
| biomarker_96 | aac(6')-IIc | dfrA19 | qnrA1.var1 | 0.56 | 0.54 | 0.1 | 38 |
| biomarker_97 | aac(6')-IIc | dfrA19 | sul1_2 | 0.56 | 0.54 | 0.1 | 38 |
| biomarker_98 | aac(6')-Ib3.var1 | aac(6')-IIc | qnrA1.var1 | 0.56 | 0.55 | 0.04 | 37 |
| biomarker_99 | blaIMP-70.var1 | catA2.var1 | qnrA1.var1 | 0.56 | 0.56 | 0.07 | 49 |
| biomarker_100 | catA2.var1 | qnrA1.var1 | sul1_2 | 0.56 | 0.56 | 0.07 | 49 |
| biomarker_101 | aac(6')-Ib3.var1 | aph(6)-Id_1 | catA2.var1 | 0.56 | 0.57 | 0.06 | 45 |
| biomarker_102 | aph(3'')-Ib_5.var1 | aph(6)-Id_1 | blaIMP-70.var1 | 0.56 | 0.57 | 0.08 | 55 |
| biomarker_103 | aph(6)-Id_1 | blaIMP-70.var1 | sul1_2 | 0.56 | 0.57 | 0.08 | 55 |
| biomarker_104 | blaIMP-70.var1 | blaTEM-1B | mcr-9 | 0.56 | 0.57 | 0.11 | 56 |
| biomarker_105 | aac(6')-IIc | aph(3'')-Ib_5.var1 | aph(6)-Id_1 | 0.56 | 0.57 | 0.02 | 39 |
| biomarker_106 | aac(6')-IIc | aph(3'')-Ib_5.var1 | blaIMP-70.var1 | 0.56 | 0.57 | 0.02 | 39 |
| biomarker_107 | aac(6')-IIc | aph(3'')-Ib_5.var1 | blaTEM-1B | 0.56 | 0.57 | 0.02 | 39 |
| biomarker_108 | aac(6')-IIc | aph(3'')-Ib_5.var1 | sul1_2 | 0.56 | 0.57 | 0.02 | 39 |
| biomarker_109 | aac(6')-IIc | aph(6)-Id_1 | blaIMP-70.var1 | 0.56 | 0.57 | 0.02 | 39 |
| biomarker_110 | aac(6')-IIc | aph(6)-Id_1 | blaTEM-1B | 0.56 | 0.57 | 0.02 | 39 |
| biomarker_111 | aac(6')-IIc | aph(6)-Id_1 | sul1_2 | 0.56 | 0.57 | 0.02 | 39 |
| biomarker_112 | aac(6')-Ib3.var1 | aph(3'')-Ib_5.var1 | aph(6)-Id_1 | 0.55 | 0.51 | 0.1 | 56 |
| biomarker_113 | aac(6')-Ib3.var1 | aph(6)-Id_1 | sul1_2 | 0.55 | 0.51 | 0.1 | 56 |
| biomarker_114 | aph(6)-Id_1 | blaIMP-70.var1 | dfrA19 | 0.55 | 0.53 | 0.07 | 54 |
| biomarker_115 | blaIMP-70.var1 | blaTEM-1B | qnrA1.var1 | 0.55 | 0.56 | 0.07 | 56 |
| biomarker_116 | blaTEM-1B | qnrA1.var1 | sul1_2 | 0.55 | 0.56 | 0.07 | 56 |
| biomarker_117 | aac(6')-Ib3.var1 | aac(6')-IIc | dfrA19 | 0.55 | 0.57 | 0.07 | 36 |
| biomarker_118 | aph(6)-Id_1 | catA2.var1 | dfrA19 | 0.55 | 0.57 | 0.07 | 48 |
| biomarker_119 | aac(6')-Ib3.var1 | blaTEM-1B | qnrA1.var1 | 0.54 | 0.53 | 0.09 | 53 |
| biomarker_120 | aac(6')-Ib3.var1 | aph(3'')-Ib_5.var1 | blaIMP-70.var1 | 0.54 | 0.54 | 0.05 | 55 |
| biomarker_121 | blaIMP-70.var1 | dfrA19 | mcr-9 | 0.54 | 0.55 | 0.07 | 56 |
| biomarker_122 | aph(6)-Id_1 | mcr-9 | qnrA1.var1 | 0.54 | 0.56 | 0.04 | 52 |
| biomarker_123 | aac(6')-Ib3.var1 | blaIMP-70.var1 | mcr-9 | 0.53 | 0.49 | 0.09 | 54 |
| biomarker_124 | aac(6')-Ib3.var1 | catA2.var1 | dfrA19 | 0.53 | 0.49 | 0.09 | 46 |
| biomarker_125 | aac(6')-Ib3.var1 | aph(6)-Id_1 | blaTEM-1B | 0.53 | 0.54 | 0.06 | 55 |
| biomarker_126 | aac(6')-Ib3.var1 | aph(6)-Id_1 | dfrA19 | 0.52 | 0.51 | 0.08 | 55 |
| biomarker_127 | aac(6')-Ib3.var1 | aac(6')-IIc | mcr-9 | 0.52 | 0.51 | 0.08 | 36 |
| biomarker_128 | aph(6)-Id_1 | blaTEM-1B | qnrA1.var1 | 0.52 | 0.52 | 0.04 | 52 |
| biomarker_129 | blaIMP-70.var1 | blaTEM-1B | dfrA19 | 0.52 | 0.56 | 0.07 | 56 |
| biomarker_130 | aac(6')-Ib3.var1 | aph(6)-Id_1 | qnrA1.var1 | 0.51 | 0.44 | 0.14 | 50 |
| biomarker_131 | aac(6')-Ib3.var1 | aph(6)-Id_1 | blaIMP-70.var1 | 0.51 | 0.5 | 0.07 | 52 |
| biomarker_132 | aac(6')-Ib3.var1 | mcr-9 | qnrA1.var1 | 0.49 | 0.45 | 0.08 | 52 |
| biomarker_133 | aac(6')-Ib3.var1 | blaIMP-70.var1 | blaTEM-1B | 0.49 | 0.47 | 0.06 | 55 |
| biomarker_134 | aac(6')-Ib3.var1 | blaIMP-70.var1 | dfrA19 | 0.49 | 0.49 | 0.04 | 54 |
| biomarker_135 | aac(6')-Ib3.var1 | aph(3'')-Ib_5.var1 | qnrA1.var1 | 0.48 | 0.51 | 0.05 | 53 |
| biomarker_136 | aac(6')-Ib3.var1 | dfrA19 | qnrA1.var1 | 0.48 | 0.51 | 0.05 | 53 |
| biomarker_137 | aph(3'')-Ib_5.var1 | blaSHV-12 | dfrA19 | 0.45 | 0.44 | 0.03 | 27 |
| biomarker_138 | aph(6)-Id_1 | blaSHV-12 | dfrA19 | 0.45 | 0.44 | 0.03 | 27 |
| biomarker_139 | blaSHV-12 | blaTEM-1B | dfrA19 | 0.45 | 0.44 | 0.03 | 27 |
| biomarker_140 | blaSHV-12 | dfrA19 | mcr-9 | 0.45 | 0.44 | 0.03 | 27 |
| biomarker_141 | blaSHV-12 | dfrA19 | sul1_2 | 0.45 | 0.44 | 0.03 | 27 |
| biomarker_142 | aac(6')-Ib3.var1 | blaSHV-12 | dfrA19 | 0.41 | 0.42 | 0.03 | 24 |
| biomarker_143 | aac(6')-Ib3.var1 | aph(3'')-Ib_5.var1 | blaSHV-12 | 0.39 | 0.35 | 0.06 | 25 |
| biomarker_144 | aac(6')-Ib3.var1 | aph(6)-Id_1 | blaSHV-12 | 0.39 | 0.35 | 0.06 | 25 |
| biomarker_145 | aac(6')-Ib3.var1 | blaSHV-12 | mcr-9 | 0.39 | 0.35 | 0.06 | 25 |
| biomarker_146 | aac(6')-Ib3.var1 | blaSHV-12 | blaTEM-1B | 0.38 | 0.38 | 0.06 | 26 |
| biomarker_147 | aac(6')-Ib3.var1 | blaSHV-12 | sul1_2 | 0.38 | 0.38 | 0.06 | 26 |
| biomarker_148 | blaSHV-12 | catA2.var1 | dfrA19 | 0.35 | 0.34 | 0.03 | 22 |
| biomarker_149 | aph(3'')-Ib_5.var1 | blaIMP-70.var1 | blaSHV-12 | 0.34 | 0.29 | 0.08 | 22 |
| biomarker_150 | aph(6)-Id_1 | blaIMP-70.var1 | blaSHV-12 | 0.34 | 0.29 | 0.08 | 22 |
| biomarker_151 | blaIMP-70.var1 | blaSHV-12 | mcr-9 | 0.34 | 0.29 | 0.08 | 22 |
| biomarker_152 | aph(3'')-Ib_5.var1 | blaSHV-12 | catA2.var1 | 0.34 | 0.33 | 0.04 | 23 |
| biomarker_153 | aph(6)-Id_1 | blaSHV-12 | catA2.var1 | 0.34 | 0.33 | 0.04 | 23 |
| biomarker_154 | blaSHV-12 | catA2.var1 | mcr-9 | 0.34 | 0.33 | 0.04 | 23 |
| biomarker_155 | blaIMP-70.var1 | blaSHV-12 | dfrA19 | 0.34 | 0.34 | 0.04 | 21 |
| biomarker_156 | blaSHV-12 | blaTEM-1B | catA2.var1 | 0.34 | 0.36 | 0.07 | 24 |
| biomarker_157 | blaSHV-12 | catA2.var1 | sul1_2 | 0.34 | 0.36 | 0.07 | 24 |
| biomarker_158 | aac(6')-Ib3.var1 | aac(6')-IIc | blaSHV-12 | 0.33 | 0.32 | 0.08 | 22 |
| biomarker_159 | aac(6')-IIc | blaIMP-70.var1 | blaSHV-12 | 0.33 | 0.32 | 0.08 | 22 |
| biomarker_160 | aac(6')-IIc | blaSHV-12 | blaTEM-1B | 0.33 | 0.32 | 0.08 | 22 |
| biomarker_161 | aac(6')-IIc | blaSHV-12 | catA2.var1 | 0.33 | 0.32 | 0.08 | 22 |
| biomarker_162 | aac(6')-IIc | blaSHV-12 | sul1_2 | 0.33 | 0.32 | 0.08 | 22 |
| biomarker_163 | aac(6')-Ib3.var1 | blaSHV-12 | qnrA1.var1 | 0.32 | 0.29 | 0.08 | 21 |
| biomarker_164 | aac(6')-IIc | blaSHV-12 | qnrA1.var1 | 0.32 | 0.29 | 0.08 | 21 |
| biomarker_165 | blaIMP-70.var1 | blaSHV-12 | qnrA1.var1 | 0.32 | 0.29 | 0.08 | 21 |
| biomarker_166 | blaSHV-12 | blaTEM-1B | qnrA1.var1 | 0.32 | 0.29 | 0.08 | 21 |
| biomarker_167 | blaSHV-12 | catA2.var1 | qnrA1.var1 | 0.32 | 0.29 | 0.08 | 21 |
| biomarker_168 | blaSHV-12 | qnrA1.var1 | sul1_2 | 0.32 | 0.29 | 0.08 | 21 |
| biomarker_169 | aac(6')-IIc | aph(3'')-Ib_5.var1 | blaSHV-12 | 0.32 | 0.34 | 0.05 | 21 |
| biomarker_170 | aac(6')-IIc | aph(6)-Id_1 | blaSHV-12 | 0.32 | 0.34 | 0.05 | 21 |
| biomarker_171 | aac(6')-IIc | blaSHV-12 | mcr-9 | 0.32 | 0.34 | 0.05 | 21 |
| biomarker_172 | aac(6')-Ib3.var1 | blaIMP-70.var1 | blaSHV-12 | 0.31 | 0.31 | 0.07 | 23 |
| biomarker_173 | aac(6')-Ib3.var1 | blaSHV-12 | catA2.var1 | 0.31 | 0.31 | 0.07 | 23 |
| biomarker_174 | blaIMP-70.var1 | blaSHV-12 | blaTEM-1B | 0.31 | 0.31 | 0.07 | 23 |
| biomarker_175 | blaIMP-70.var1 | blaSHV-12 | catA2.var1 | 0.31 | 0.31 | 0.07 | 23 |
| biomarker_176 | blaIMP-70.var1 | blaSHV-12 | sul1_2 | 0.31 | 0.31 | 0.07 | 23 |
| biomarker_177 | aac(6')-IIc | blaSHV-12 | dfrA19 | 0.29 | 0.29 | 0.04 | 20 |
| biomarker_178 | aph(3'')-Ib_5.var1 | blaSHV-12 | qnrA1.var1 | 0.29 | 0.29 | 0.04 | 20 |
| biomarker_179 | aph(6)-Id_1 | blaSHV-12 | qnrA1.var1 | 0.29 | 0.29 | 0.04 | 20 |
| biomarker_180 | blaSHV-12 | dfrA19 | qnrA1.var1 | 0.29 | 0.29 | 0.04 | 20 |
| biomarker_181 | blaSHV-12 | mcr-9 | qnrA1.var1 | 0.29 | 0.29 | 0.04 | 20 |

### 2.2 Plasmid analysis

**Figure S2.**
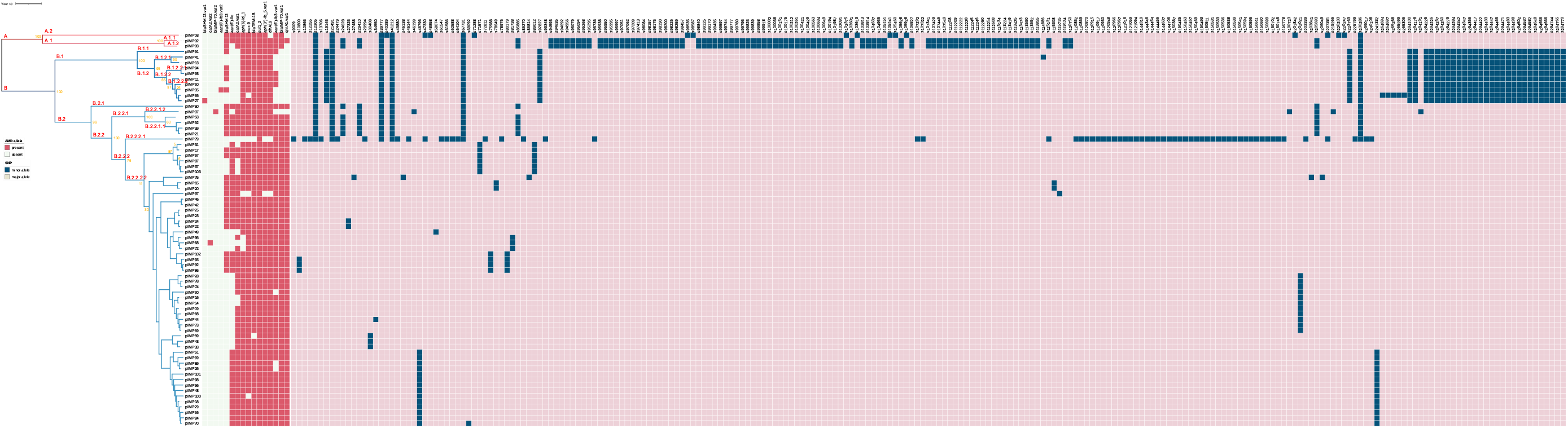
Alignment between lineages, AMR alleles, and core-genome single-nucleotide polymorphisms (SNPs) of IncHI2 plasmids. Lineages of 72 IncHI2 plasmids are labelled (red, with bootstrap values shown aside in gold) and coloured (only for major lineages) in a Bayesian dated phylogenetic tree of the plasmids^2^. The red heat map shows allelic presence-absence of acquired AMR genes that are likely to be carried by these plasmids according to a previous study^6^. The blue heat map shows core-genome SNPs of the plasmids when compared them to a reference plasmid genome pKA_P10 (RefSeq accession: NZ_CP044215.1). Coordinates of the SNPs in the reference genome are used as column names (with a prefix ‘s’) for the heat map. Zoom in to show labels.

### 2.3 Relationship between genomic variation and spatial-temporal proximity

We also investigated whether our spatial-temporal proximity aligned with transmission by comparing core-genome SNP distances between IncHI2 plasmids identified in CPE_IMP_ isolates from corresponding patients (Figure S3). We would expect higher patient similarities (indicating a higher probability of transmission) with lower genomic diversity (lower number of SNPs). Conversely, higher genomic diversity (lower number of SNPs) between patients would not support evidence of transmission. We found, the optimised contact network *G*_*m*_ recovered edges were skewed towards low genomic diversity, reflecting that our model tends to pick edges representing a higher likelihood of transmission (Figure S3)A. Moreover, we found a negative correlation between *G*_*m*_ recovered edges and genomic diversity, meaning that as our *G*_*m*_ indicates of a lower probability of transmission, the genomic diversity increases between the plasmid of a patients CPE_IMP_, also less indicative of transmission.

**Figure S3.**
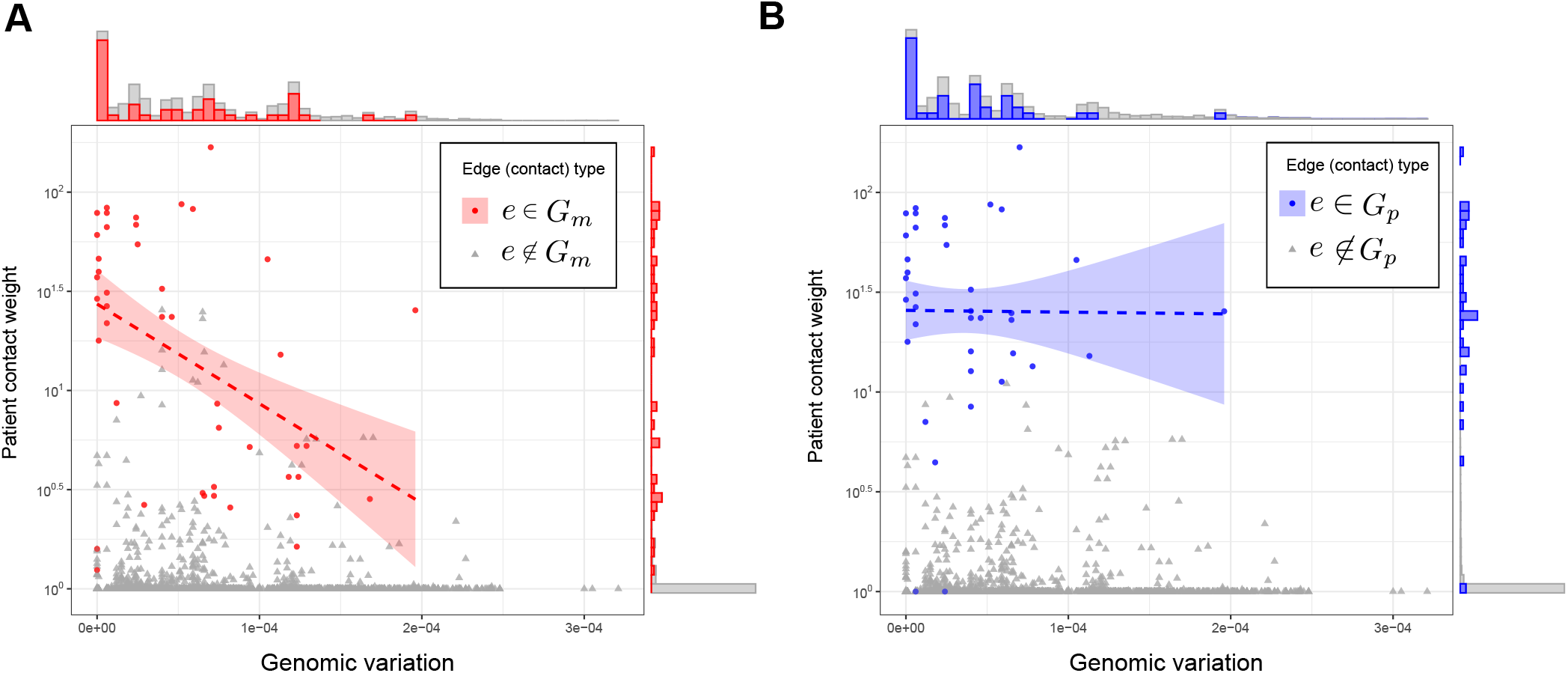
Comparison of genomics variation and StEP proximity. Panels show the SNP distances between IncHI2 plasmids obtained from WGS patient CPE_IMP_ isolates, and the StEP contact proximity weights. Since not all patients had WGS, not all comparisons are possible, and hence only edges for which both patients had WGS are visualised in the figure. Panel **A**: comparison with edges (contacts) highlighted which were selected in StEP’s contact network *G*_*m*_. Panel **B**: contacts edges in a network of physical contact from *G*_*p*_.

Looking specifically at edges selected in the optimised contact network *G*_*m*_, there is a visible heterogeneity in the chosen weights. Most edges from *G*_*m*_ have high weightings and are visibly distinct from the total edge space, several (few) lower weighted edges are also selected. Our models’ heterogeneous selection is a desirable effect missed by standard network construction approaches given the topological plain, non-uniform, and occurring over several different periods of time. Overall, the total edge space in terms of patient movement similarity is skewed, indicating most are unlikely to result in transmission - lining up with only a single edge needed to capture transmission. Compared to edges resulting from physical contacts in *G*_*p*_ (Figure S3B) there is considerable overlap with 64 physical edges recovered in *G*_*m*_. However, there is no decreasing relationship between patient similarity weighting and genomic variation, as observed in (Figure S3)A. This difference suggests that, first, not all physical contacts are conclusive of transmission; and second, in order to optimally characterise the transmission, some physical proximity is redundant (24 not included *G*_*m*_).

### 2.4 Topological comparison analysis

**Figure S4.**
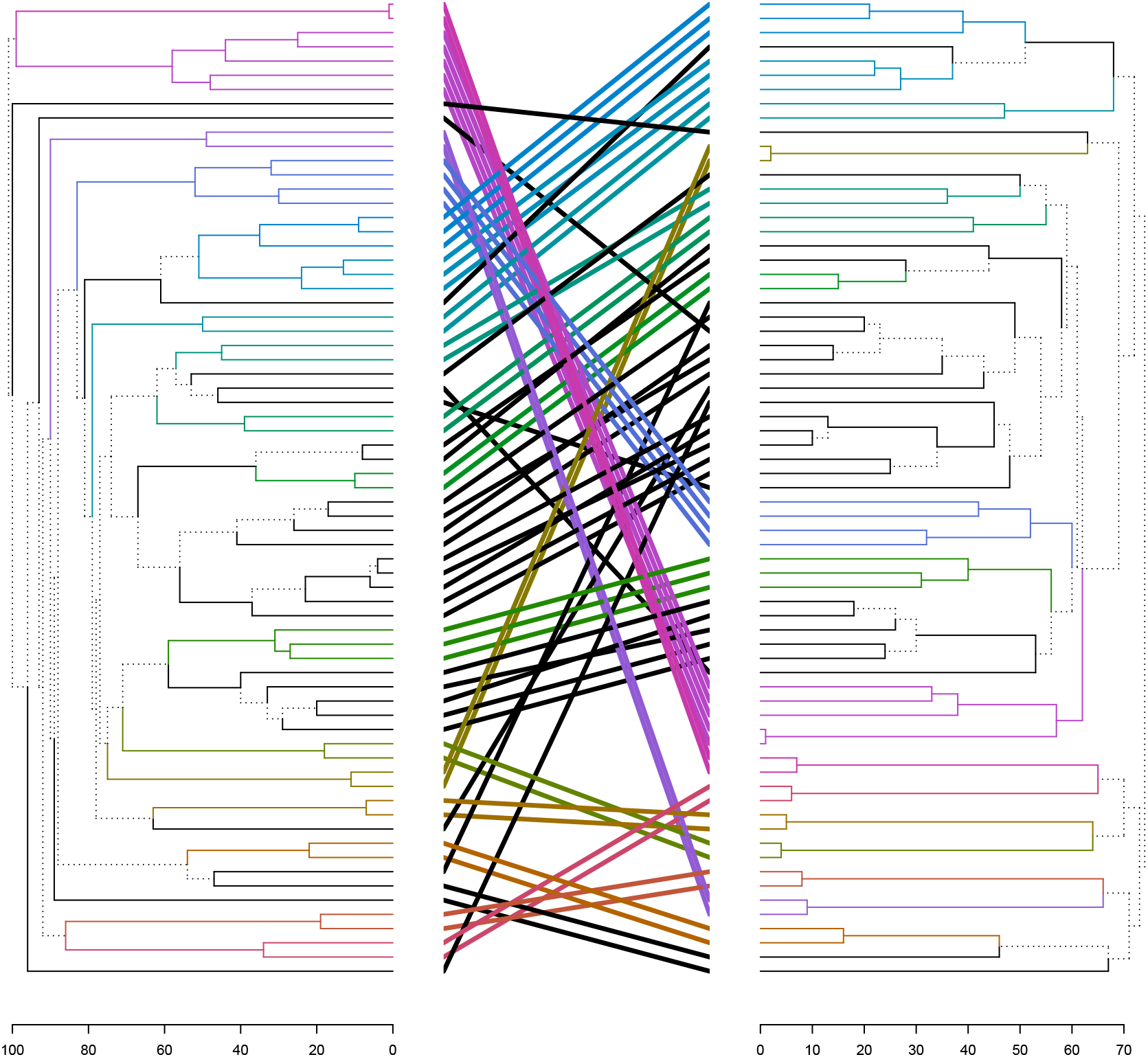
Tangle-gram comparing model derived contact network *G*_*m*_ (left) to physical contact network *G*_*p*_ (right). Both networks were hierarchically clustered using walktrap^7^ and aligned with dendextend^8^. Distinct branches (not shared between clustering) are marked with a dashed line, whereas shared branches are solid and colour coded according by corresponding branches. Remove node names, add graph labels, make distinct lines dotted.

**Figure S5.**
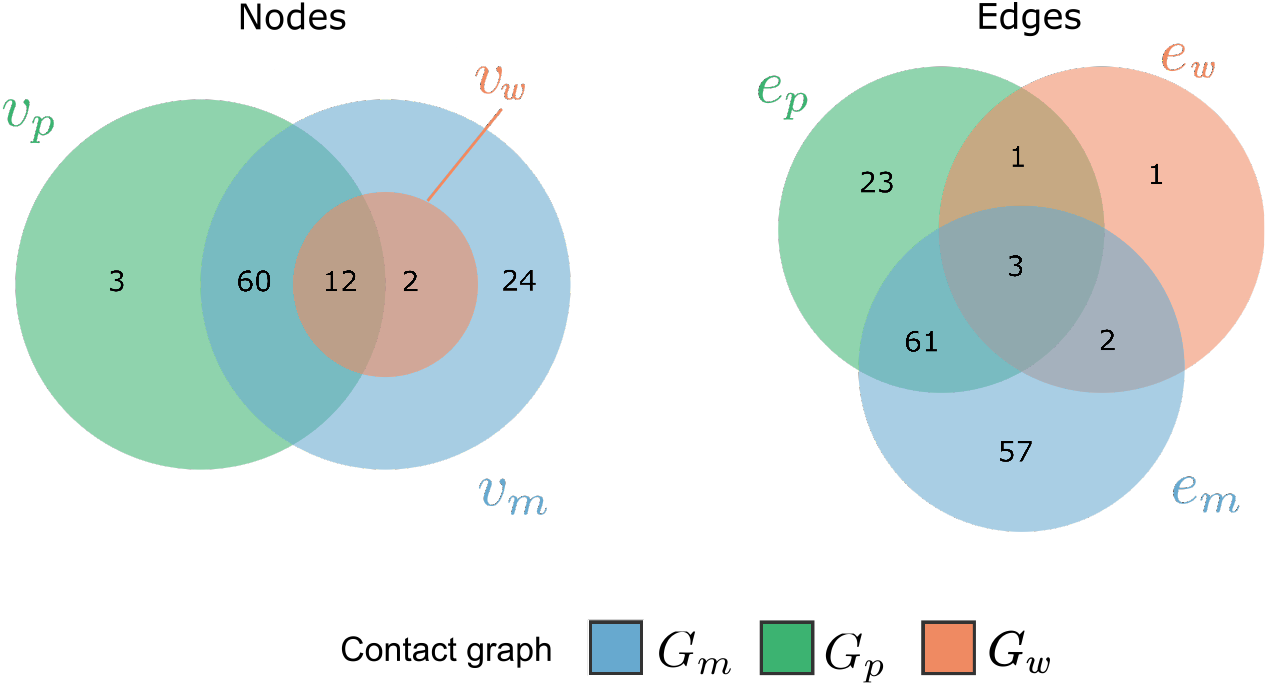
Venn diagram for overlap in nodes and edges between CPE_IMP_ contact networks: (1) Model recovered contact graph *G*_*m*_ which captures indirect contact through background patient movement, (2) Physical contact graph *G*_*p*_, capturing contact between patients which were on the same ward on the same day, and (3) contacts from the result of a standard outbreak investigation *G*_*w*_ which had only looked at contact between patients on the ward where they were identified as positive (+/- 7 days).

### 2.5 Topological signals from StEP’s contact network

**Figure S6.**
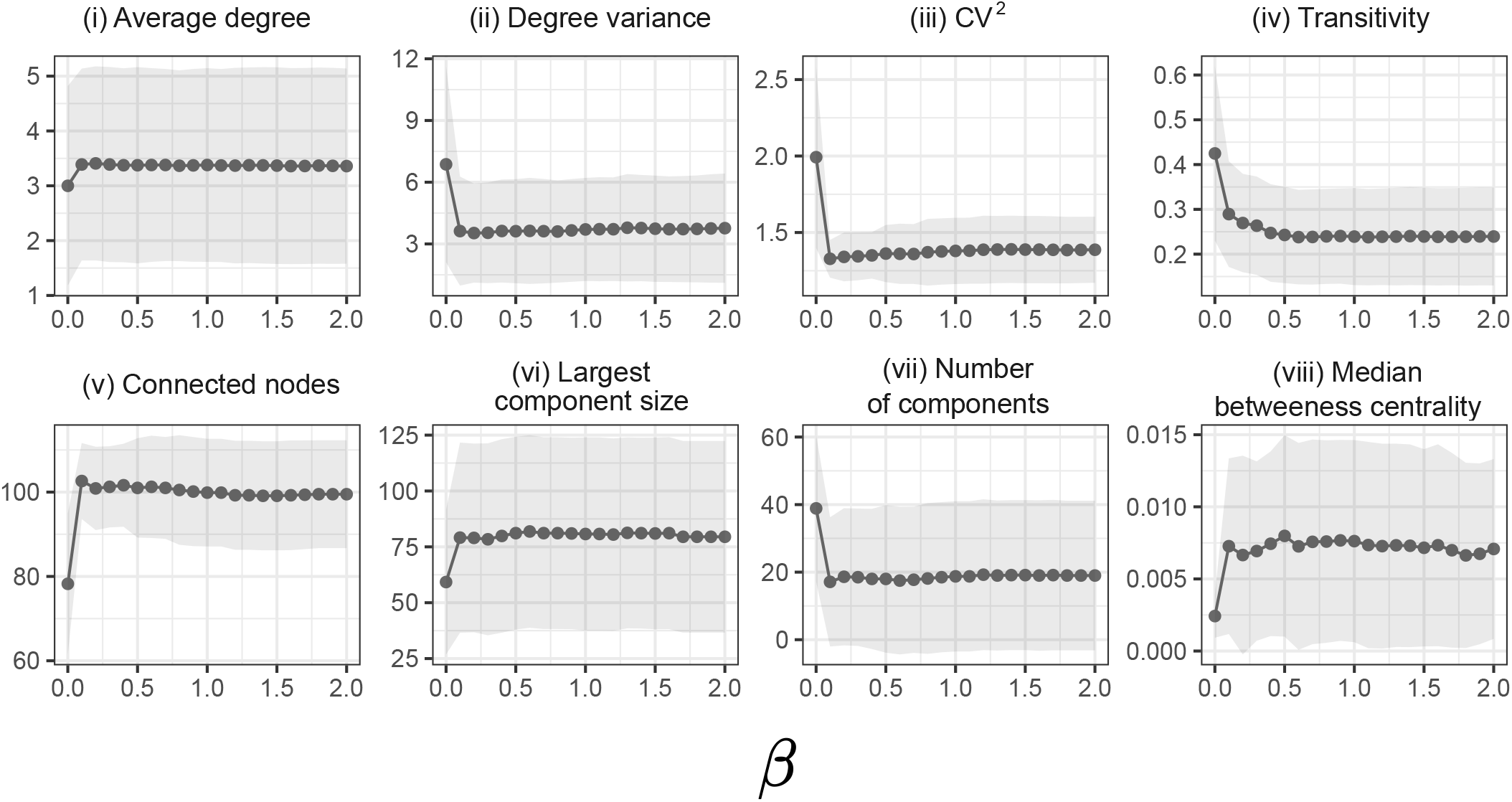
Measures of network topology as a function of propagation speed *β* for StEP’s CPE_IMP_ contact network. Metrics: **(i)** average degree, **(ii)** degree variance, **(iii)** CV^2^, **(iv)** transitivity, **(v)** connected nodes, **(vi)** largest component size, **(vii)** number of components, **(viii)** and median betweenness centrality. As opposed to variation by edge density *k*, propagation speed *β* exhibits largely less influence over network metrics, except initially when *β* = 0.

**Figure S7.**
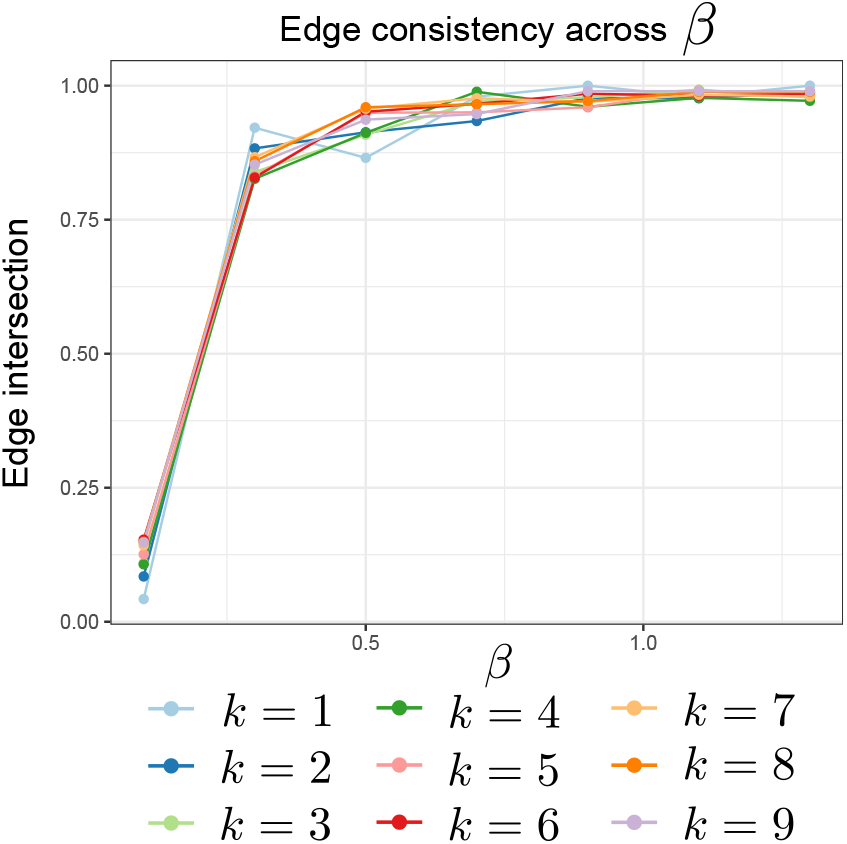
Network edge consistency as a function of propagation speed *β*. For the two model parameters, propagation speed *β* and edge density *k*, we look at the edge consistency across parameter values (measured as the proportion of edges intersecting between two edge sets, |*E* ∩ *E*^′^| */* | *E* ∪ *E*^′^|). For lower values in propagation speed, varying the parameter has a large impact on changes in edges. However, for propagation speed *β >* 0.5 varying the parameter has little effect on the edges recovered, with edges remaining greater than 90% consistent.

## 3 Case study 2

### 3.1 Contact network comparison

**Figure S8.**
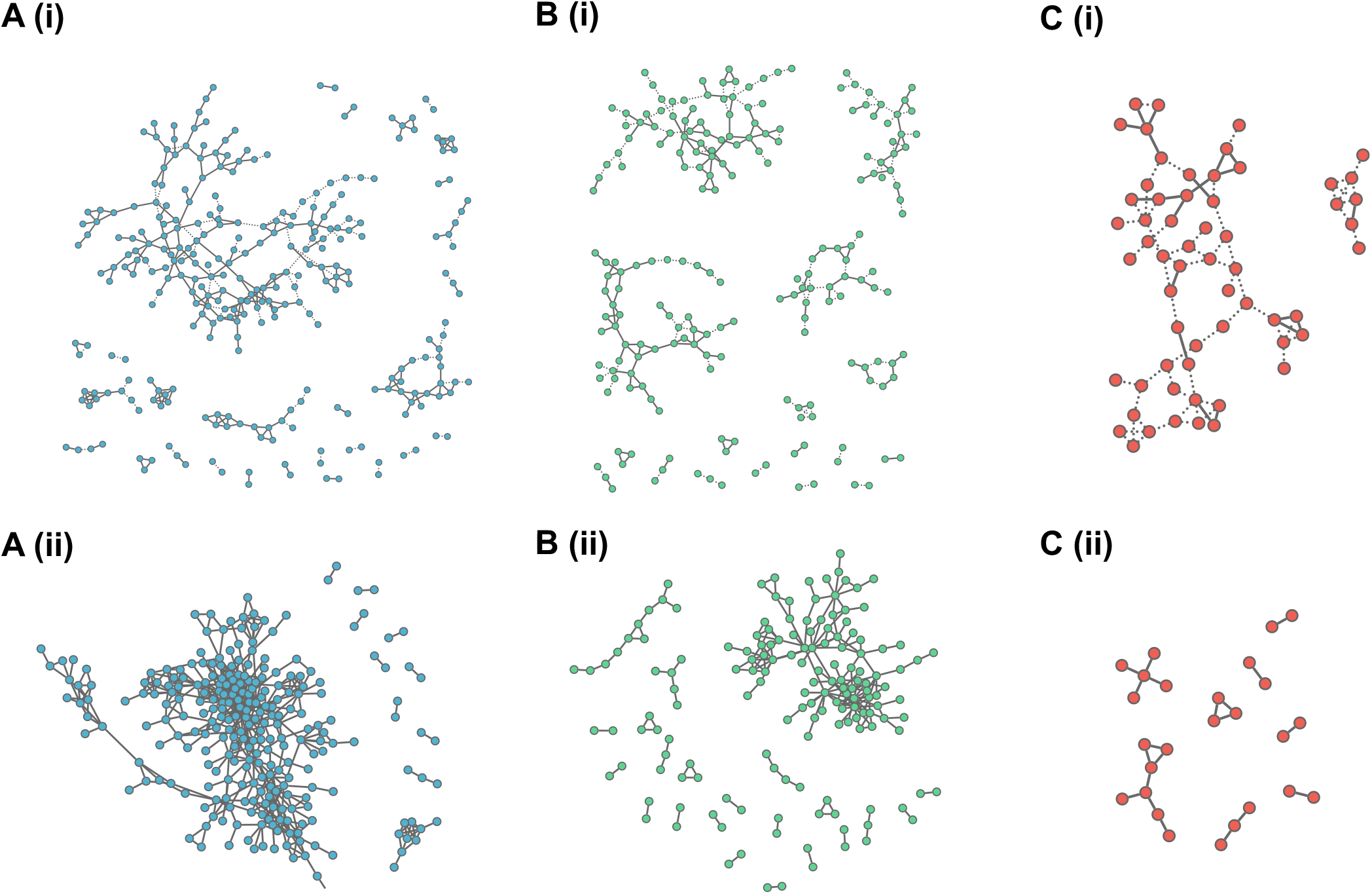
Contact network models for CPE_OXA-48_, CPE_NDM_, and CPE_VIM_. Panels **A-C** shows the contact networks for CPE_OXA-48_, CPE_NDM_, and CPE_VIM_ and respectively. Panels **A (i)**,**B (i), C (i)** correspond to StEP’s recovered contact networks *G*_*m*_ (with edge density *k* = 3, and propagation speed *β* = 0.6), whereas panels **A (ii)**,**B (ii), C (ii)** are the contact networks constructed from physical contact alone (*G*_*p*_). Across all CPE types StEP links a greater number of patients (CPE_OXA-48_: 382 versus 269, CPE_NDM_: 201 versus 156, CPE_VIM_: 59 versus 26, Table S2). Connected cases in the physical contact network largely overlapped with those identified by StEP (88.8%, 94.9%, and 100% for CPE_OXA-48_, CPE_NDM_, and CPE_VIM_ respectively).

### 3.2 Quantifying alignment to bacterial species

**Figure S9.**
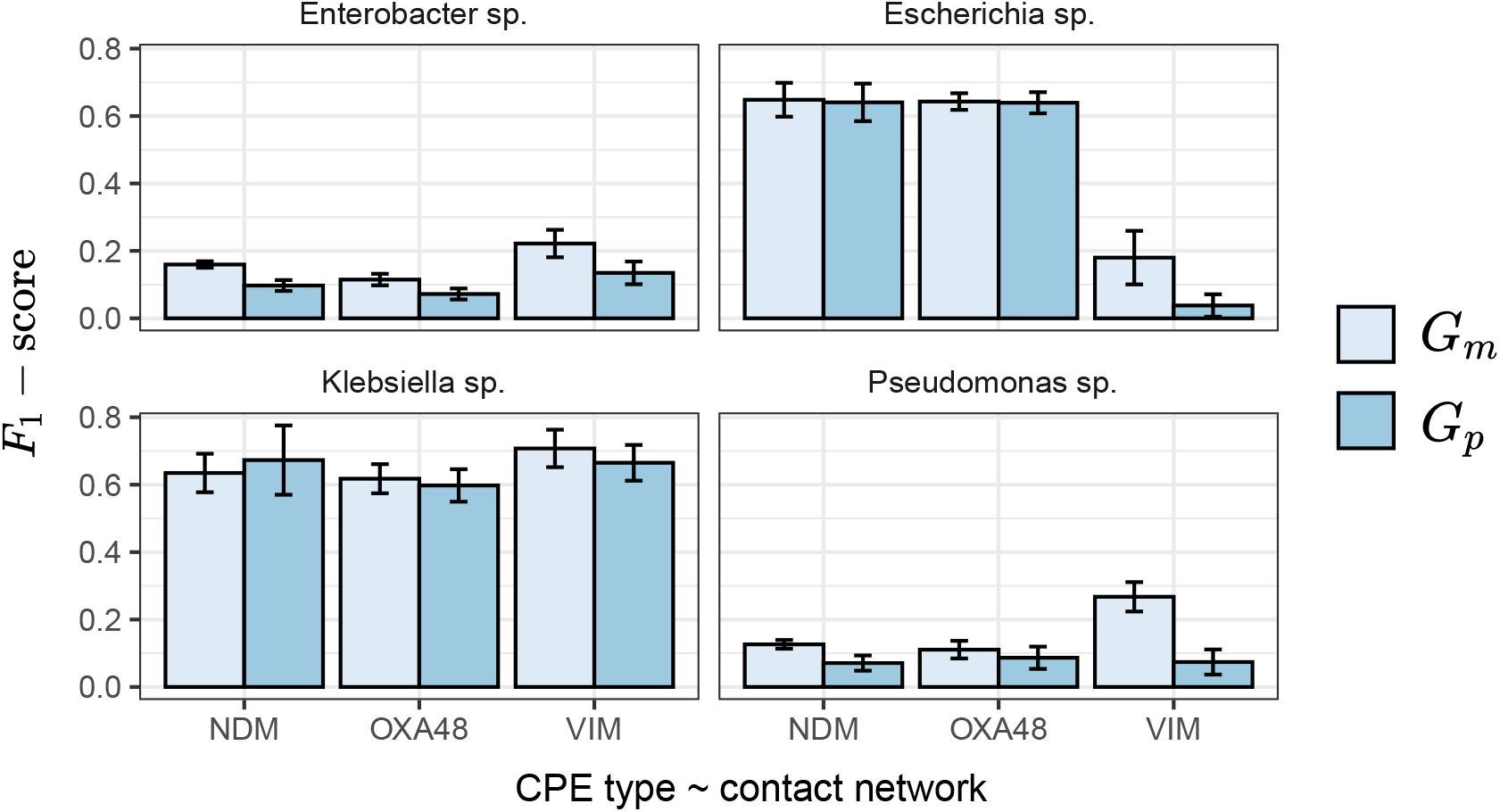
Alignment of CPE type contact network to bacterial species. We computed the alignment between each CPE contact network and bacterial species GDR (determined using the *F*_1_-score from GDR (see Methods: GDR and Methods: Classification metric)) where node labels are bacterial species. In this framework a higher *F*_1_-score would suggest that movement of bacterial species between individuals is well captured via the contact structure. Whilst the *Enterbactor sp*. and the Pseudomonas sp. seem to be little captured by the contact networks, across all CPE types, *G*_*m*_ produces a better alignment to the distrubtion of bacterial species (exhibited by the higher *F*_1_-score). Additionaly, both *Escherichia sp*. and Klebsiella sp. aligned largely to both contact networks, but most so towards *G*_*m*_.

### 3.3 Topological comparison analysis

**Figure S10.**
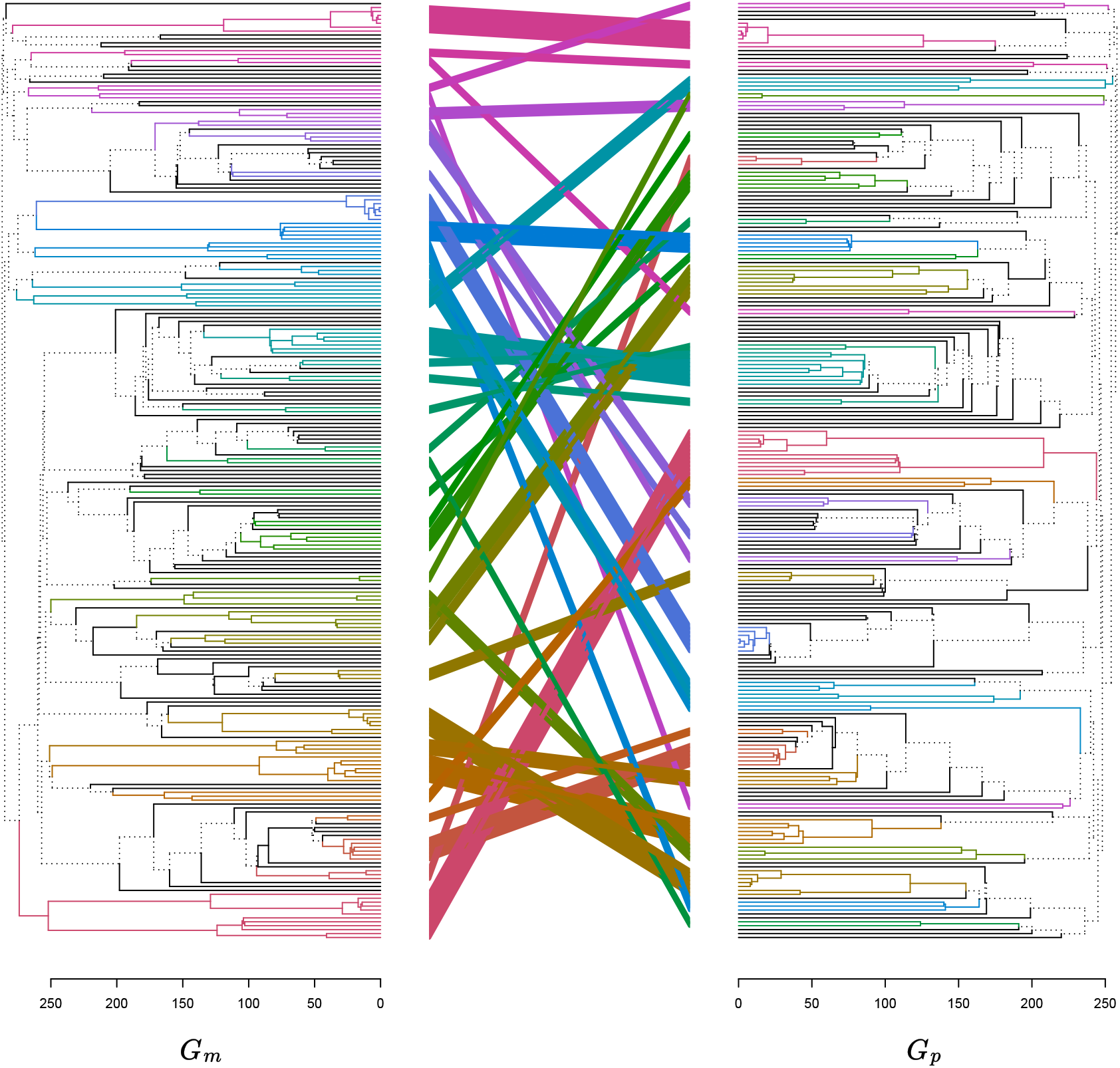
Tangle-gram comparing CPE_OXA-48_ model derived contact network (left) to physical contact network. Both networks were hierarchically clustered using walktrap^7^ and aligned with dendextend^8^. Distinct branches (not shared between clustering) are marked with a dashed line, whereas shared branches are solid and colour coded according by corresponding branches.

**Figure S11.**
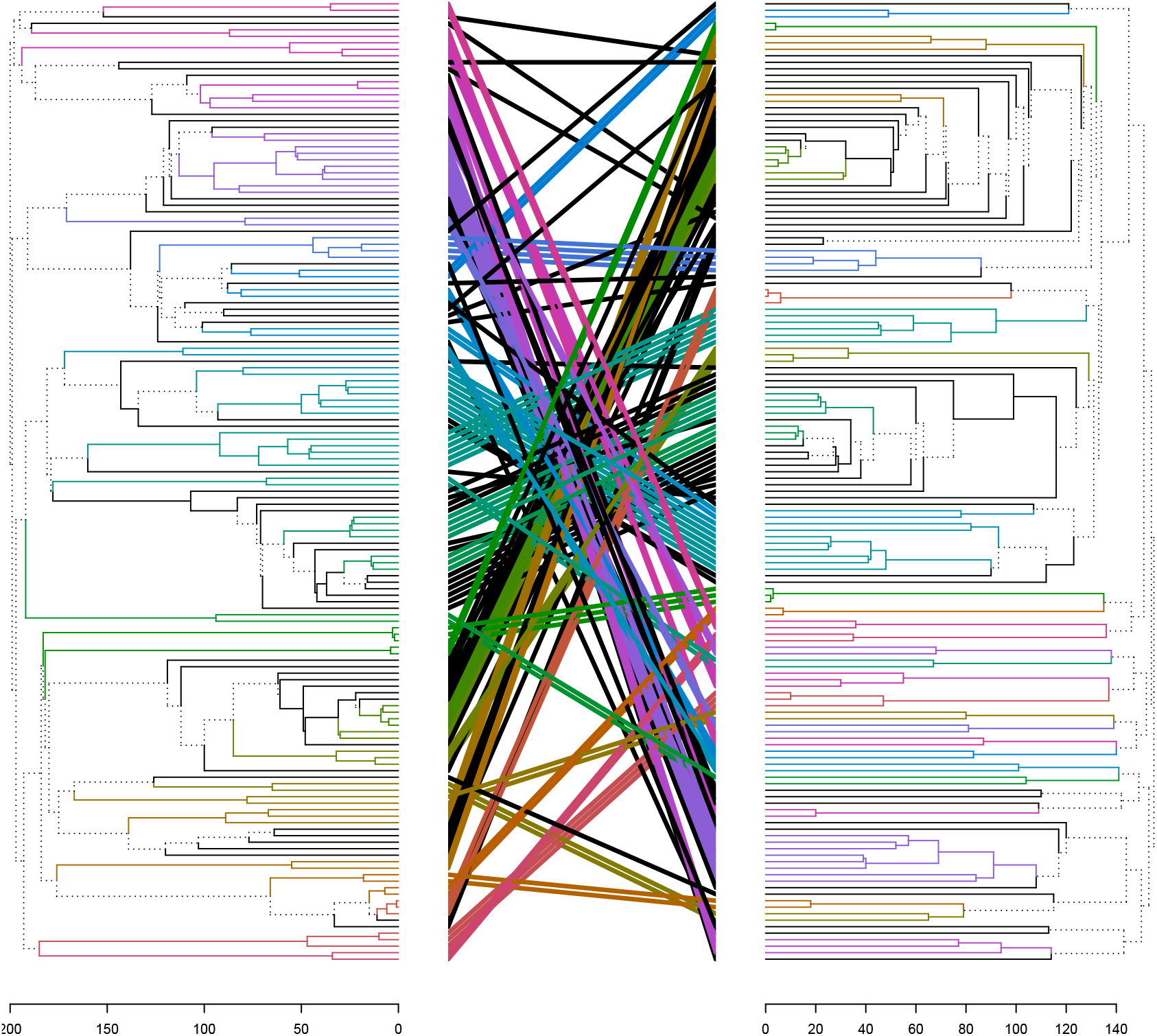
Tangle-gram comparing CPE_NDM_ model derived contact network (left) to physical contact network. Both networks were hierarchically clustered using walktrap^7^ and aligned with dendextend^8^. Distinct branches (not shared between clustering) are marked with a dashed line, whereas shared branches are solid and colour coded according by corresponding branches.

**Figure S12.**
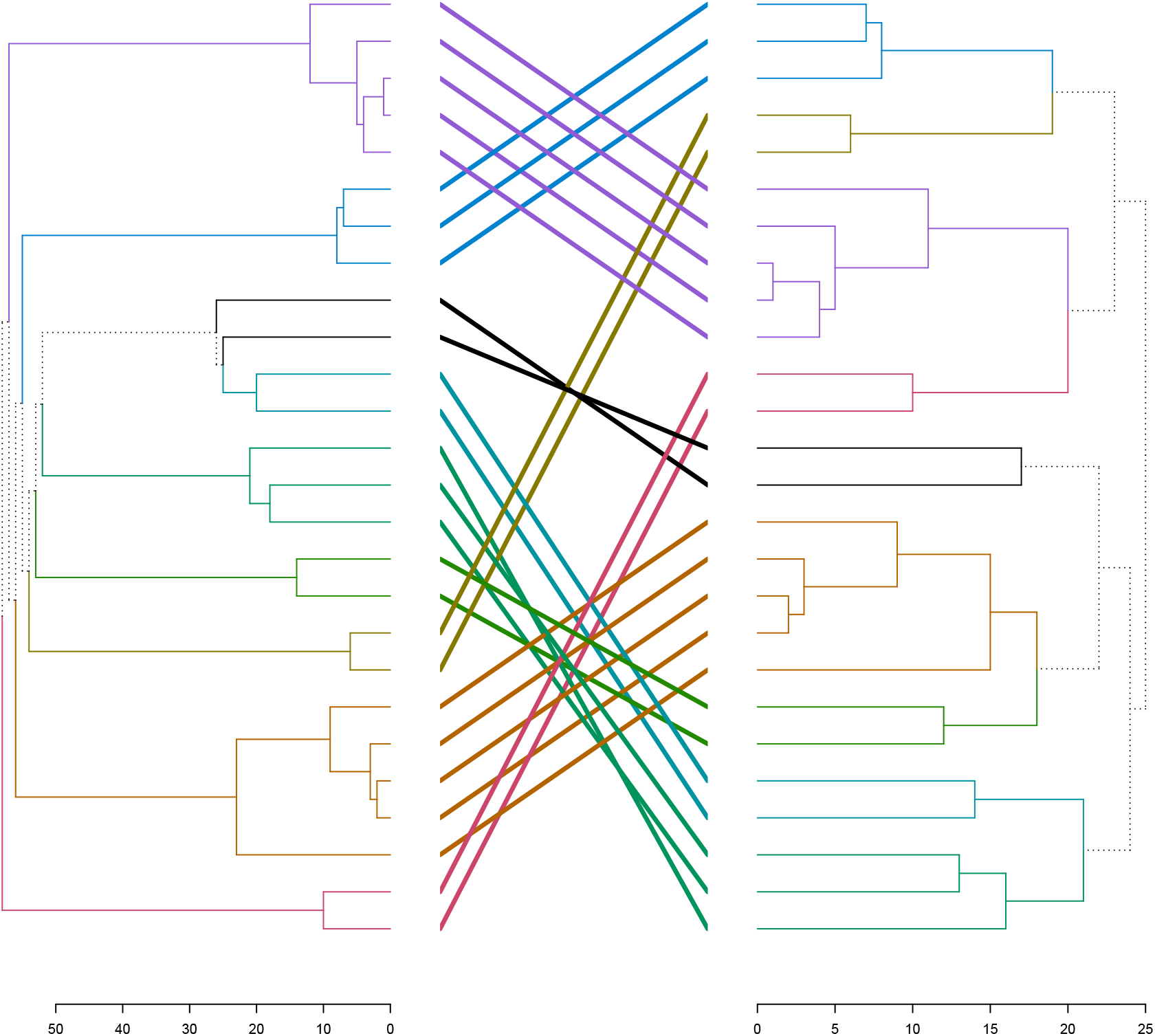
Tangle-gram comparing CPE_VIM_ model derived contact network (left) to physical contact network. Both networks were hierarchically clustered using walktrap^7^ and aligned with dendextend^8^. Distinct branches (not shared between clustering) are marked with a dashed line, whereas shared branches are solid and colour coded according by corresponding branches.

**Figure S13.**
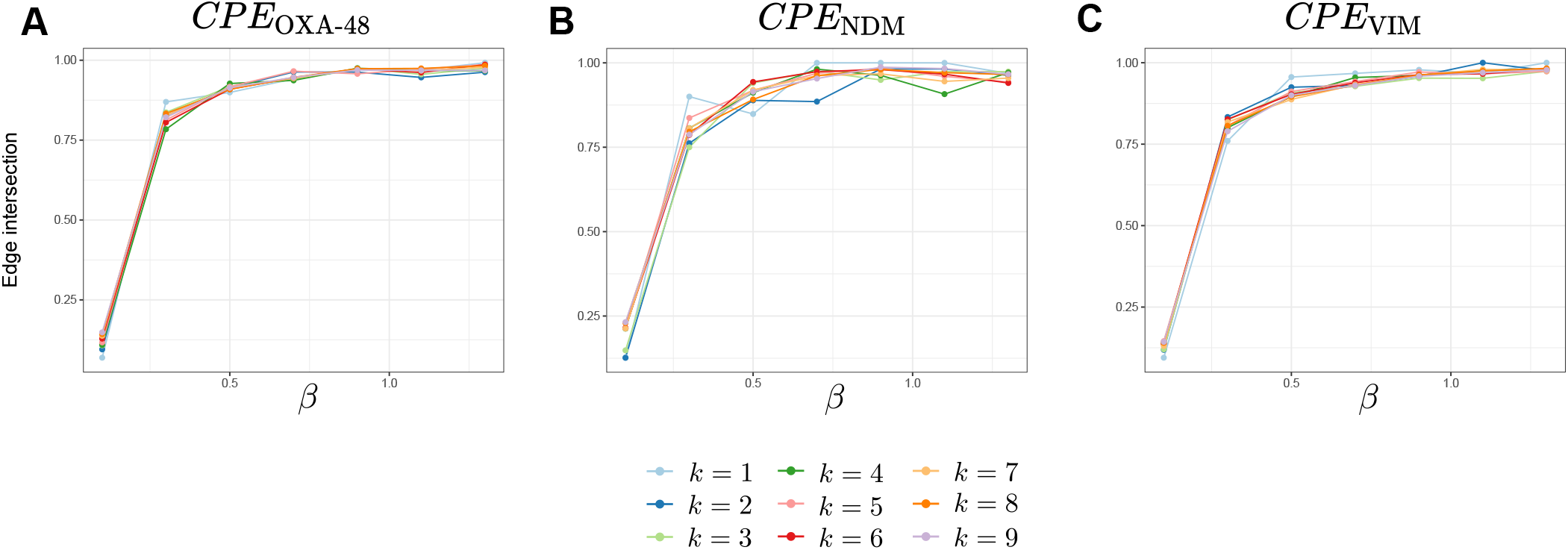
Network edge consistency as a function of propagation speed *β* for CPE contact networks. For the two model parameters, propagation speed *β* and edge density *k*, we look at the edge consistency across parameter values (measured as the proportion of edges intersecting between two edge sets, | *E*∩ *E*^′^ | */* | *E*∪ *E*^′^ |). Across all CPE contact networks, for lower values in propagation speed, varying the parameter has a large impact on changes in edges. However, for propagation speed *β >* 0.5 varying the parameter has little effect on the edges recovered, with edges remaining greater than 90% consistent.

**Figure S14.**
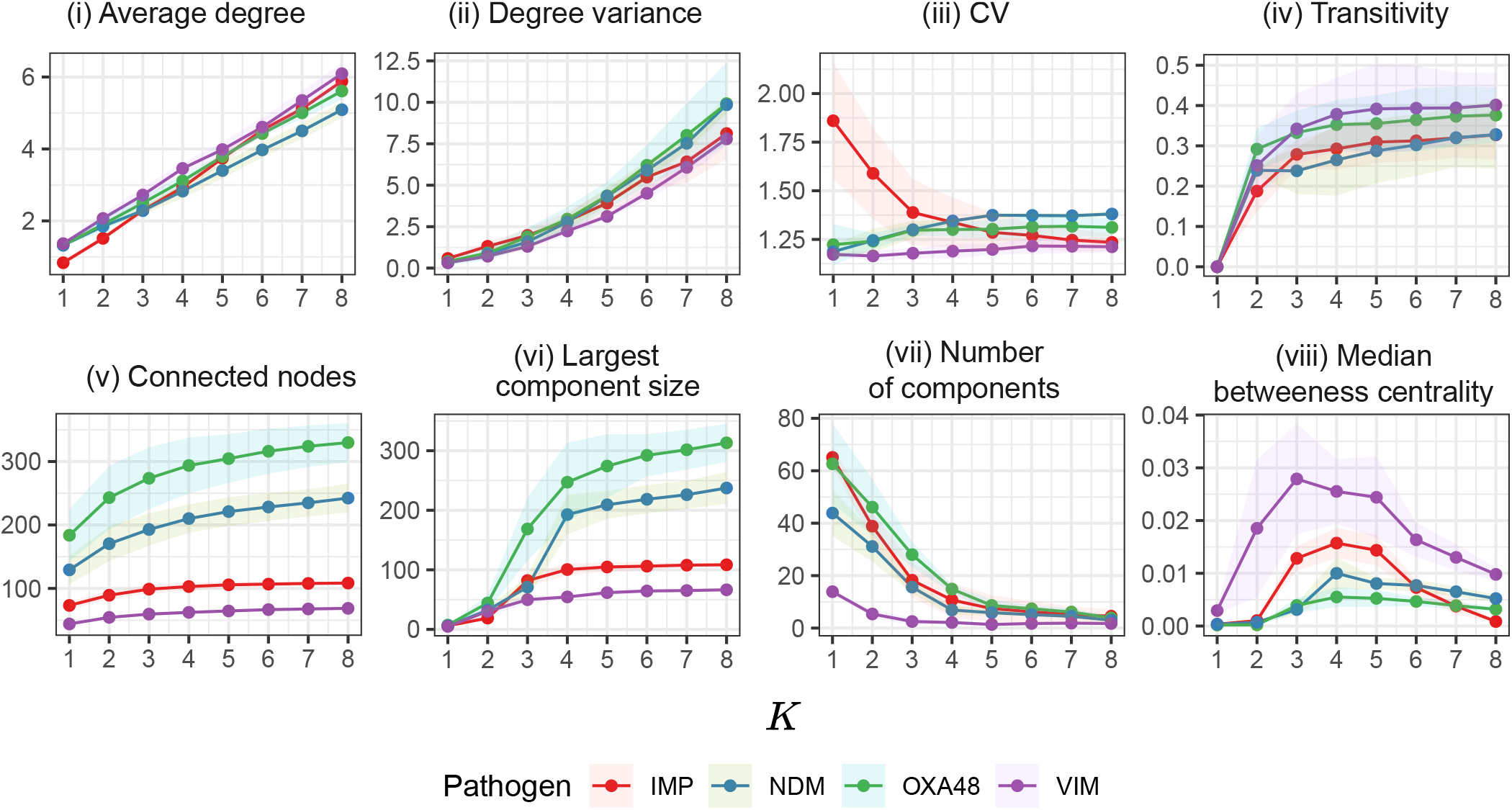
Measures of network topology as a function of edge density *k* comparing StEP’s CPE contact networks. Metrics: **(i)** average degree, **(ii)** degree variance, **(iii)** CV^2^, **(iv)** transitivity, **(v)** connected nodes, **(vi)** largest component size, **(vii)** number of components, **(viii)** and median betweenness centrality.

**Figure S15.**
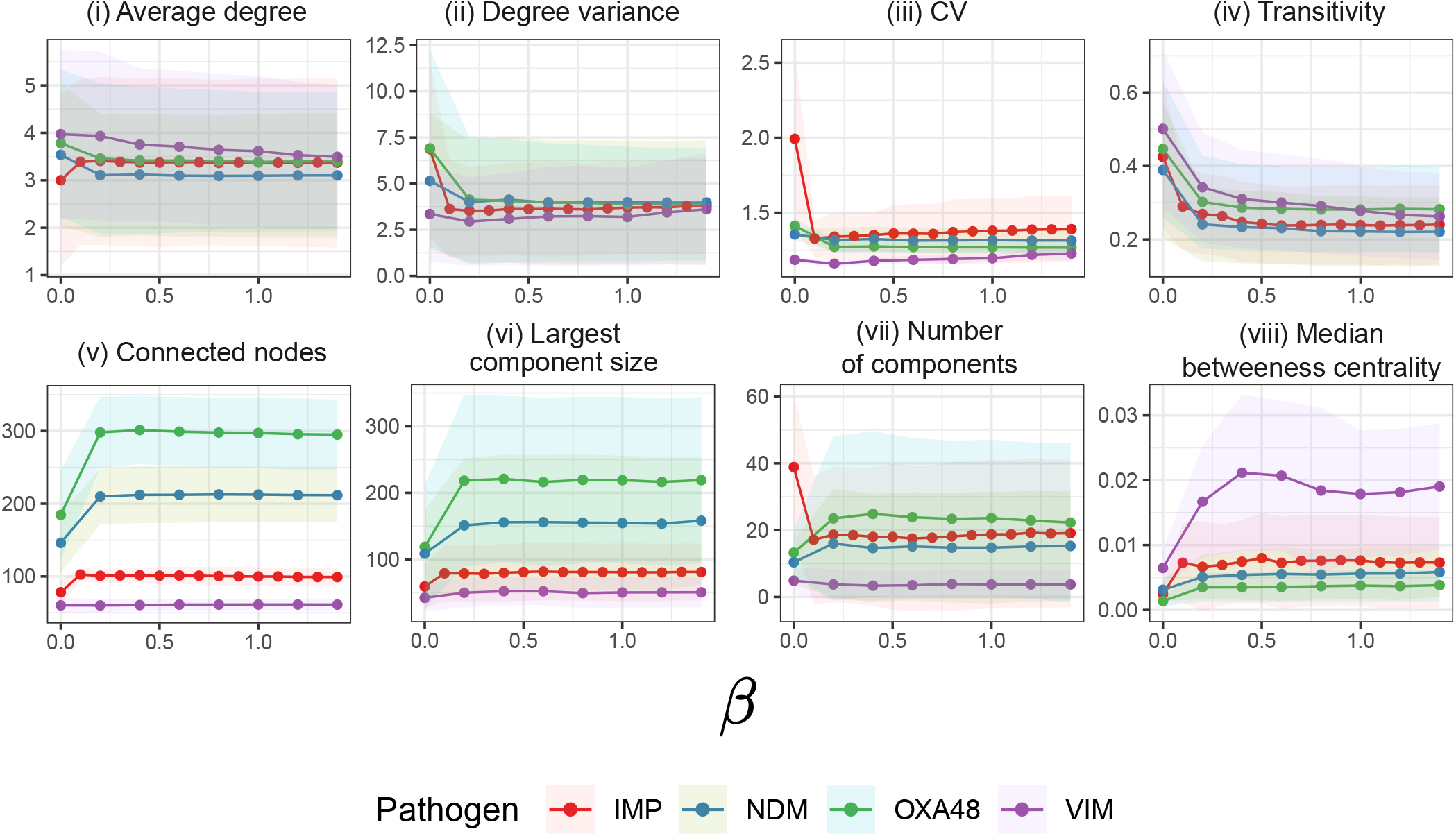
Measures of network topology as a function of propergation speed *β* comparing StEP’s CPE contact networks. Metrics: **(i)** average degree, **(ii)** degree variance, **(iii)** CV^2^, **(iv)** transitivity, **(v)** connected nodes, **(vi)** largest component size, **(vii)** number of components, **(viii)** and median betweenness centrality.

**Table S2.** Edge density *k* elbows detected for the CPE_OXA-48_, CPE_NDM_, and CPE_VIM_ contact networks. Metrics examined: **(i)** average degree, **(ii)** degree variance, **(iii)** CV^2^, **(iv)** transitivity, **(v)** connected nodes, **(vi)** largest component size, **(vii)** number of components, **(viii)** and median betweenness centrality.

| Graph | $CV^2$ | Transitivity | Connected nodes | Largest component | Number of components | Median betweenness centrality | Average |
| --- | --- | --- | --- | --- | --- | --- | --- |
| $CPE_{OXA-48}$ | 3 | 2 | 4 | 4 | 5 | 4 | 3.7 |
| $CPE_{NDM}$ | 5 | 2 | 4 | 4 | 4 | 4 | 3.8 |
| $CPE_{VIM}$ | - | 3 | 3 | 3 | 3 | 3 | 3 |
| Average | 4 | 2.3 | 3.7 | 3.7 | 4 | 3.7 | 3.5 |

## 4 Case study 3

### 4.1 Topological comparison analysis

**Figure S16.**
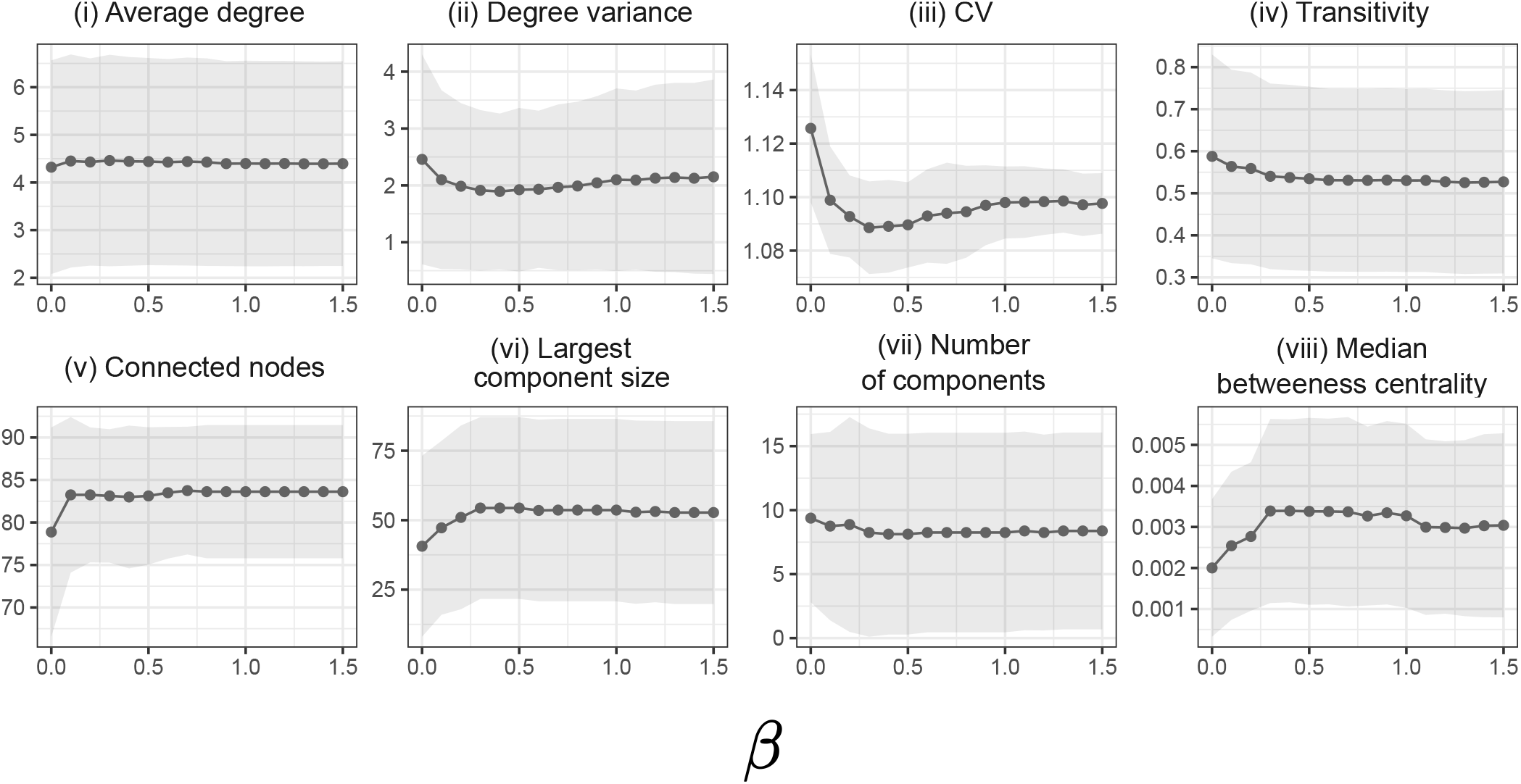
Measures of network topology as a function of propagation speed *β*. Metrics: **(i)** average degree, **(ii)** degree variance, **(iii)** CV^2^, **(iv)** transitivity, **(v)** connected nodes, **(vi)** largest component size, **(vii)** number of components, **(viii)** and median betweenness centrality.

**Figure S17.**
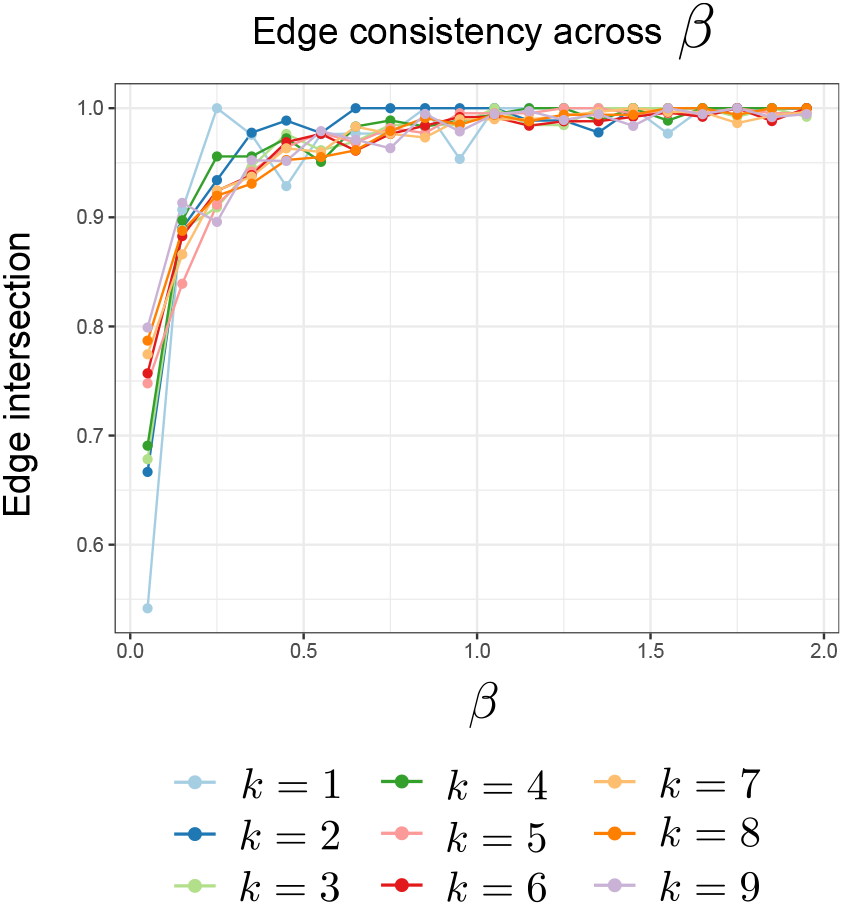
Network edge consistency as a function of propagation speed *β*. For the two model parameters, propagation speed *β* and edge density *k*, we look at the edge consistency across parameter values (measured as the proportion of edges intersecting between two edge sets, |*E* ∩ *E*^′^| */*| *E*∪ *E*^′^|). For lower values in propagation speed, varying the parameter has a large impact on changes in edges. However, for propagation speed *β >* 0.5 varying the parameter has little effect on the edges recovered, with edges remaining greater than 90% consistent.

**Figure S18.**
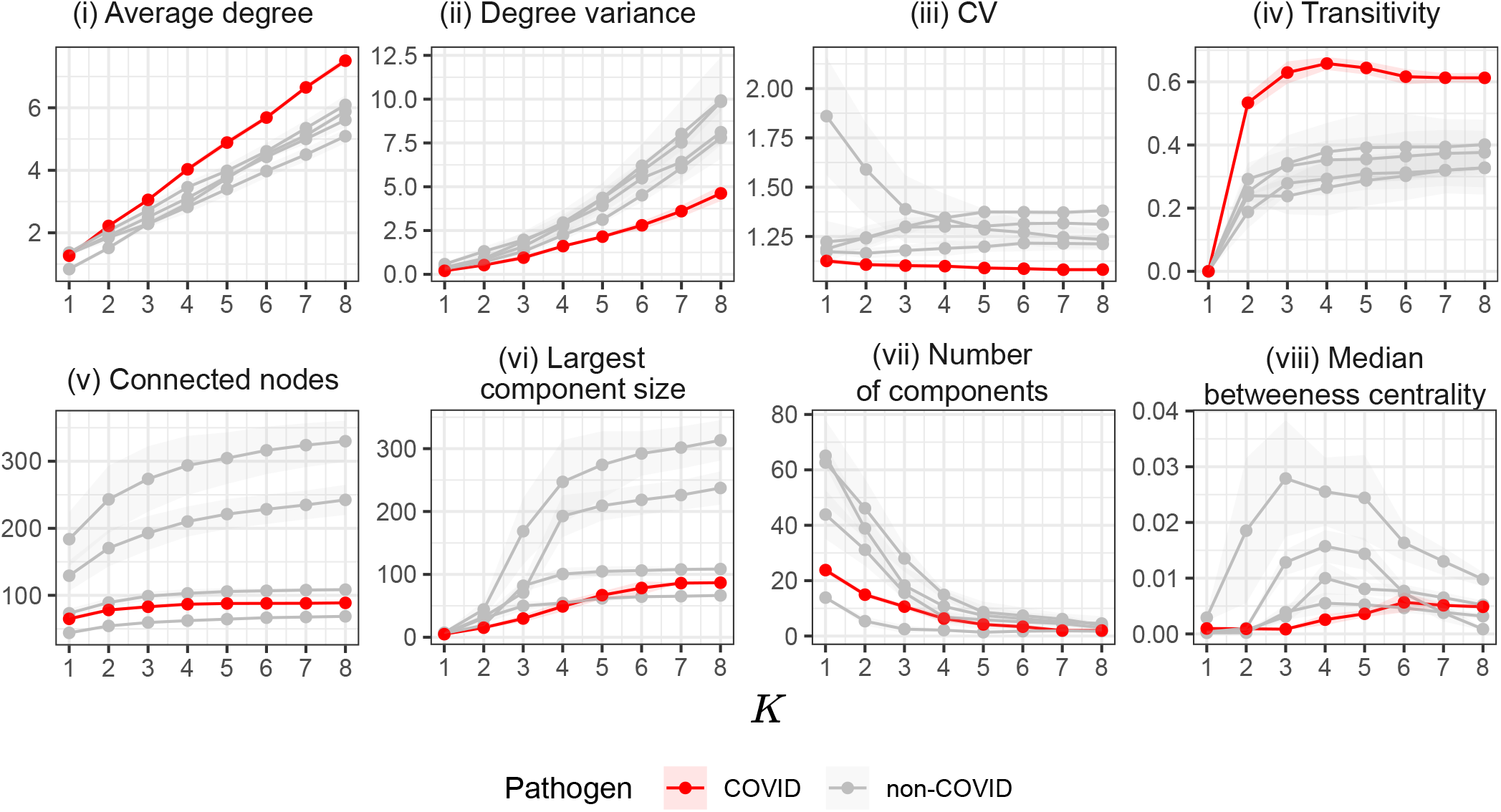
Measures of network topology as a function of edge density *k* comparing StEP’s COVID contact network to the CPE. Metrics: **(i)** average degree, **(ii)** degree variance, **(iii)** CV^2^, **(iv)** transitivity, **(v)** connected nodes, **(vi)** largest component size, **(vii)** number of components, **(viii)** and median betweenness centrality.

